# A deep cellular atlas of the human ventral substantia nigra in Parkinson’s identifies a genetic and molecular overlap with insulin resistance

**DOI:** 10.1101/2025.05.28.25328401

**Authors:** Viola Volpato, David A. Menassa, Preethi Sheshadri, Stefania Giussani, Michal Rokicki, Lucia F. Cardo, Marirena Bafaloukou, Ann-Kathryn Schalkamp, Agata Zaremba, Jimena Monzón-Sandoval, Ngoc-Nga Vinh, Joanne Morgan, Michele T.M. Hu, Scott Miners, Richard Wade-Martins, Cynthia Sandor, Laura Parkkinen, Caleb Webber

## Abstract

Parkinson’s disease (PD) is a complex neurodegenerative disorder characterised by selective neuronal loss. We integrate deep full-length single-nuclei sequencing of the human substantia nigra with novel genome-wide association studies (GWAS) identifying genetic and cellular drivers of PD. Genetic risk converges on AGTR1+ dopaminergic neurons and perineuronal oligodendrocytes (pODCs), both reduced in PD, as well as oligodendrocyte precursor cells, enriched among disease-disrupted intercellular interactions. AGTR1+ neurons represent a metabolically stressed state, characterised by renin-angiotensin system (RAS) and MAPK activation, oxidative stress, and mitochondrial dysfunction, rather than a distinct subtype. AGTR1+ neurons and pODCs link PD risk to metabolic traits; in pODCs, this association reflects insulin resistance with downregulated PI3K–AKT signalling. GWAS of comorbid PD and type 2 diabetes (T2D) identifies loci in AGTR1 and TCF7L2, while AGTR1+ neurons specifically upregulate RAS and T2D drug targets. Familial PD genes associated to comorbid PD/T2D associate with non-Lewy body PD, stratifying disease mechanisms.

---

Parkinson’s disease (PD) is the second most common neurodegenerative disorder, affecting approximately 2-3% of individuals over the age of 65^1,2^. PD is defined pathologically by the progressive degeneration of dopaminergic neurons in the substantia nigra pars compacta (SNpc). Anatomically, the SNpc is subdivided into a dorsal tier (dorso-medial region) and a ventral tier (ventro-lateral region) of dopaminergic neurons. These tiers are considered functionally distinct populations: for example, ventral tier neurons tend to project to the dorsal striatum (motor-related regions) while dorsal tier neurons project more to ventral/medial striatal regions involved in associative or limbic functions^3^. Importantly, the degeneration in PD is not uniform across the SNpc: post-mortem analyses have shown a selective loss of neurons in the ventrolateral (ventral tier) SNpc^4,5^, particularly those expressing angiotensin receptor 1 (AGTR1) as recently reported^6^. Ventral SNpc dopaminergic neurons also preferentially express the transcription factor SOX6, along with a suite of genes related to mitochondrial respiration, suggesting ventral neurons have a higher metabolic rate and may experience more oxidative stress^7^. This neuronal loss is also accompanied by dysfunction in other cell types, including oligodendrocytes and astrocytes^8,9,10^, that altogether lead to neurological, cognitive, and motor impairments^11^. To date, only one human cellular atlas study has focussed specifically on the ventral nigra in PD but only upon the dopaminergic neuronal populations, leaving the role of ventral nigral glia in PD unclear^12,13,6^.

Epidemiological evidence links metabolic disorders, such as Type 2 Diabetes (T2D), with increased PD risk suggesting shared pathogenic pathways^14,15^. Over 50% PD patients are reported to have insulin resistance^16^ which is related to more severe phenotype and more rapid disease progression^17^. Given the high costs and lengthy timelines associated with traditional drug development, there is growing interest in repurposing existing drugs approved to treat T2D, including statins^18,19^, phosphodiesterase inhibitors^20^, and GLP1 receptor (GLP1R) agonists^21,22^, as promising candidates for PD treatment^23^. Other metabotropic drugs, such as angiotensin receptor blockers (ARBs), commonly prescribed for hypertension, have also shown neuroprotective effects in preclinical PD models, including reducing oxidative damage and inflammation^24,25^. Thus, elucidating the mechanistic interplay between selective neuronal susceptibility and metabolic dysregulation could substantially advance therapeutic strategies leveraging these existing pharmacological agents for PD treatment.

Advancements in single-cell RNA sequencing (scRNA-seq) have revolutionized our ability to analyse gene expression at the individual cell level, enabling the identification of cell-type-specific changes, molecular states, and cellular interactions in the brain. Gene expression atlases of the human SN have revealed key cellular vulnerabilities and pathways involved in PD pathology ^10,12,13,26,27,28^ but the full complexity of multi-cellular network interactions underlying disease progression remains incompletely understood. For instance, how dopaminergic neuron degeneration dynamically reshapes glial responses and whether glial activation exacerbates or mitigates neuronal loss is still unresolved.

Here, we perform deep and full length SMARTseq single-nuclei transcriptome analysis of 40 ventral SNpc samples across PD Braak stages, identifying AGTR1+ DaNs, perineuronal satellite oligodendrocytes (psODCs), oligodendrocyte precursor cells (OPCs), and reactive astrocytes as key cell types linked to PD genetic risk. We confirm in the same cases using histological methods a significant loss of AGTR1 transcripts and protein and a loss of psODCs which are secondary to SNpc neuronal loss. Our analysis reveals novel intercellular mechanisms, including PD genetic risk associated ligand-receptor interactions, beyond neuronal cell-autonomous factors. Gene isoform shifts in OPC-to-ODC differentiation correlate with a reduction in psODCs in PD. We identify AGTR1+ DaNs as the most vulnerable cell type, showing a transition to a high-metabolic-stress state marked by elevated dopamine synthesis, renin-angiotensin system (RAS) and MAPK activation, explaining the neuroprotective effects of ARBs and GLP-1R agonists. We demonstrate that T2D genetic risk is directly linked to PD through AGTR1 with genome-wide significant SNPs identified within AGTR1 gene in PD patients with comorbid T2D.

## Results

### Deep single nuclei sequencing of the human postmortem ventral SNpc

Using SmartSeq technology (Methods), we deeply sequenced the full-length transcriptomic profiles of 23,885 nuclei from postmortem ventral SNpc brain region of 20 donors (from two replicate punctures each) (Fig S1a), including 6 idiopathic PD patients at Braak stage 5-6, 3 PD patients at Braak stage 3-4, 2 cases with incidental Lewy body disease (iLBD) at Braak stage 3-4, 2 cases with iLBD at Braak stage 1-2 and 7 healthy controls with an average age of 83 years (Table S1). Quality control analyses yielded a total of 23,034 high-quality nuclei with an average of 1194 nuclei per donor (Methods). Each nucleus was deeply sequenced with an average of 351,886 total number of reads detecting an average of 8,653 expressed genes and of 115,656 transcripts per nucleus (Fig. 1c, Methods), providing approximately four times higher resolution of gene expression than previous studies ^10,12,13^. Based on the expression of marker genes we identified seven major cell types that were contributed to by all donors (Fig. 1a, Fig. 1d). We found that, while cell type proportions were less similar between donors, replicate punctures showed much higher similarity within donor (Fig. S2). These major cell types included (i) DaNs (LMX1A/B, GFRA2, GRIK3, Fig 1b) with four subtypes: DaN_0 (GRIA3, DCX), DaN_1 showing high similarity to DaN_3 in expression of marker genes (SLC6A3, GFRA2, RET, PITX3) except for AGTR1 that was specifically more expressed in DaN_3, DaN_2 marked by expression of TMEFF2(Fig 1e, Fig 1f), (ii) oligodendrocytes (ODCs) (MPBP, MOG, Fig 1b) with three subtypes: ODC_0 (PLXDC2, PALM2, LAMA2) expressing genes associated with neuronal supporting functions^29^ (Table S2), ODC_1 (PLP1, SPARC, OPALIN, PMP2, HHIP) expressing genes associated with myelin and immune response^29^ (Table S2), ODC_2 (RBFOX1, AFF3, FMN1, PDGFRA/B, PDGFC, CLDN10) expressing genes associated with perineuronal satellite oligodendrocyte (psODC_2) population involved in neuronal homeostasis and support^30^ (Fig 1e, Fig 1f), (iii) astrocytes (AQP4, GFAP, Fig 1b) with four subtypes: Astrocyte_0 (GRM5, PPFIA2, PAK3) expressing BBB-related genes^29^, Astrocyte_1 (GABRA2, EDNRB, SLC6A11) and Astrocyte_2 (PTPRT, APHA6, EPHB1, ADGRV1) expressing genes associated with neuronal supporting functions^29^ (Table S2), Astrocyte_3 (SPOCK1, MYO1E, SLC24A2, PLP1, S100B) expressing genes related to myelin^29^ (Table S2)(Fig 1e, Fig 1f), (iv) microglia (CSF1R) with three subtypes: Microglia_0 (INPP5D, ITGAM, ADAP2, VAV1) annotated as macrophage, Microglia_1 (PTPRC, GPNMB, SPP1, TYROBP, TREM2, MS4A4A) expressing genes characteristic of activated microglia, Microglia_2 (MRC1, ABCC4, TFRC, DOCK8, KCNQ1) marked by expression of macrophage-specific genes with anti-inflammatory role^31^ (Fig 1e, Fig 1f), (v) GABAergic neurons (GAD1, GAD2, ALDH1A1, Fig 1b), (vi) oligodendrocyte precursor cells (OPCs) (VCAN, Fig 1b) and (vii) T-cells (CD8A, PTPRC, Fig 1b).

**Figure 1.**
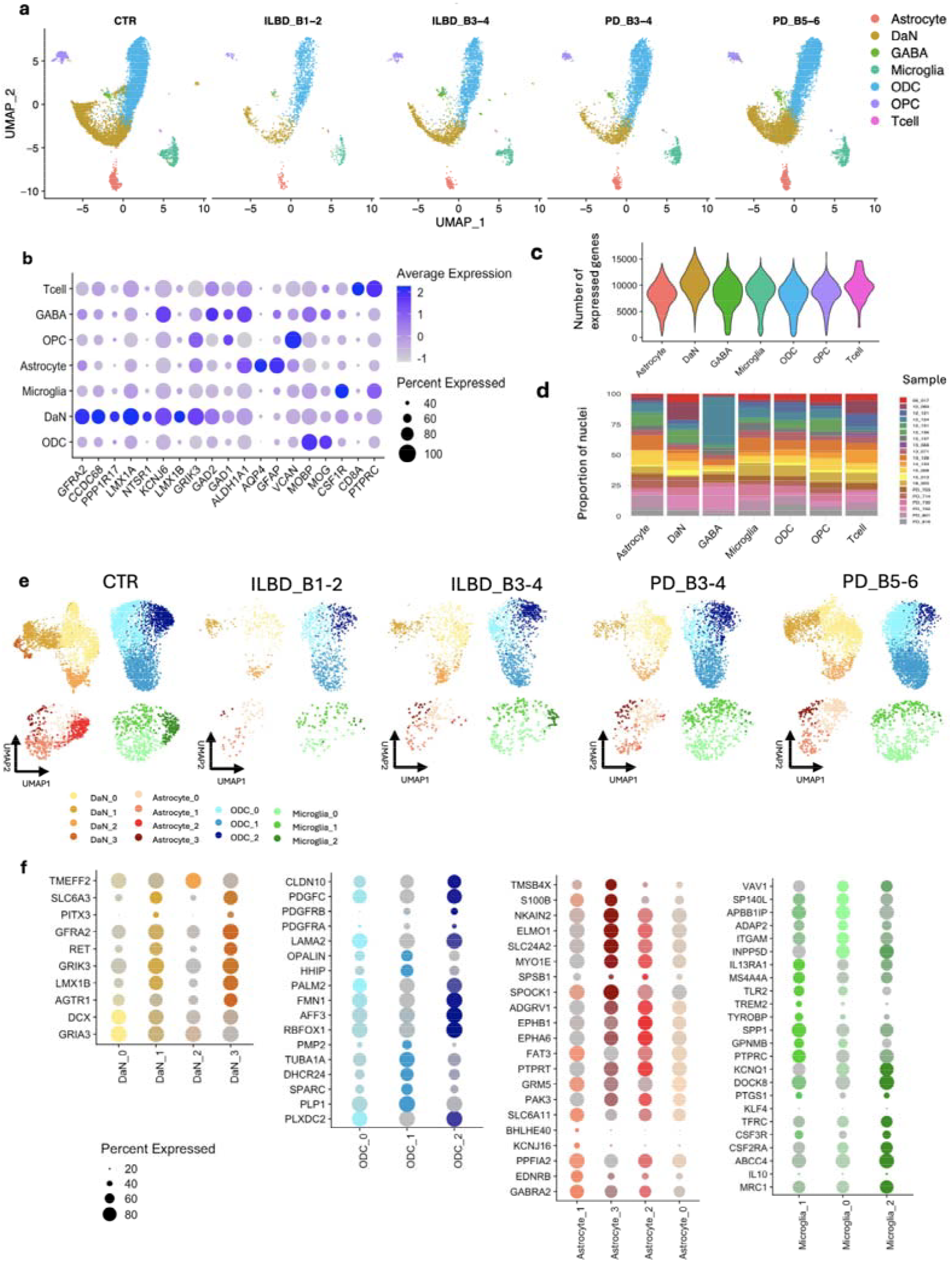
A deep single-nuclei transcriptomic atlas of the human substantia nigra. **a)** UMAP of 23,034 high quality nuclei from postmortem substantia nigra labelled according to cell type and split by disease condition. **b)** Dotplot of top markers for main seven cell types. The size of the dots represents the percentage of cells with expression. The color of the dots represents the scaled average expression level. **c)** Violin plots showing the number of protein coding genes detected per nuclei by cell type. **d)** Proportion of nuclei from each donor by cell type. **e)** UMAP of 23,034 high quality nuclei from postmortem substantia nigra labelled according to cell subtype and split by disease condition. **f)** Dotplot of top markers for cell subtypes. The size of the dots represents the percentage of cells with expression. Grey color indicates low gene expression level.

### PD genetic risk is associated with AGTR1+ DaNs, perineuronal satellite ODCs and OPCs

We first examined the enrichment of common variant risk of sporadic PD within cell-type-specific genes across ventral SNpc major cell types through MAGMA (Methods) and observed significant enrichment in OPCs (Bonferroni-corrected P<0.05) and in DaNs (nominal P<0.05)(Fig. 2a, Table S3) as previously reported^10,32^. Conditional analysis revealed that PD risk was associated with distinct sets of DaNs- and OPC-specific genes (Fig. 2a). Among cell subtypes, significant enrichments of PD heritable risk was found among cell-type-specific genes of AGTR1+ DaN_3 (Bonferroni-corrected P<0.05) in agreement with recent analyses of mouse and human single-cell datasets^6^, in DaN_1 (nominal P<0.05) and in psODC_2 (Bonferroni-corrected P<0.05) (Fig. 2b, Table S3). We found that the fraction of PD genetic risk contributing to the psODC_2 association was distinct to that fraction associated with AGTR1+ DaN_3 and vice versa (Fig. S3). The fraction of risk associated to the OPCs was also distinct to those associated with AGTR1+ DaN_3 and psODC_2 and vice versa (Fig. S3). However, the PD risk association with DaN_1 was lost once conditioned upon AGTR1+ DaN_3 expression but not lost in AGTR1+ DaN_3 when conditioned upon DaN_1 (Fig. S3) suggesting that DaN_1 genes underlying this association are a subset of those underlying the stronger AGTR1+ DaN_3 association.

**Figure 2.**
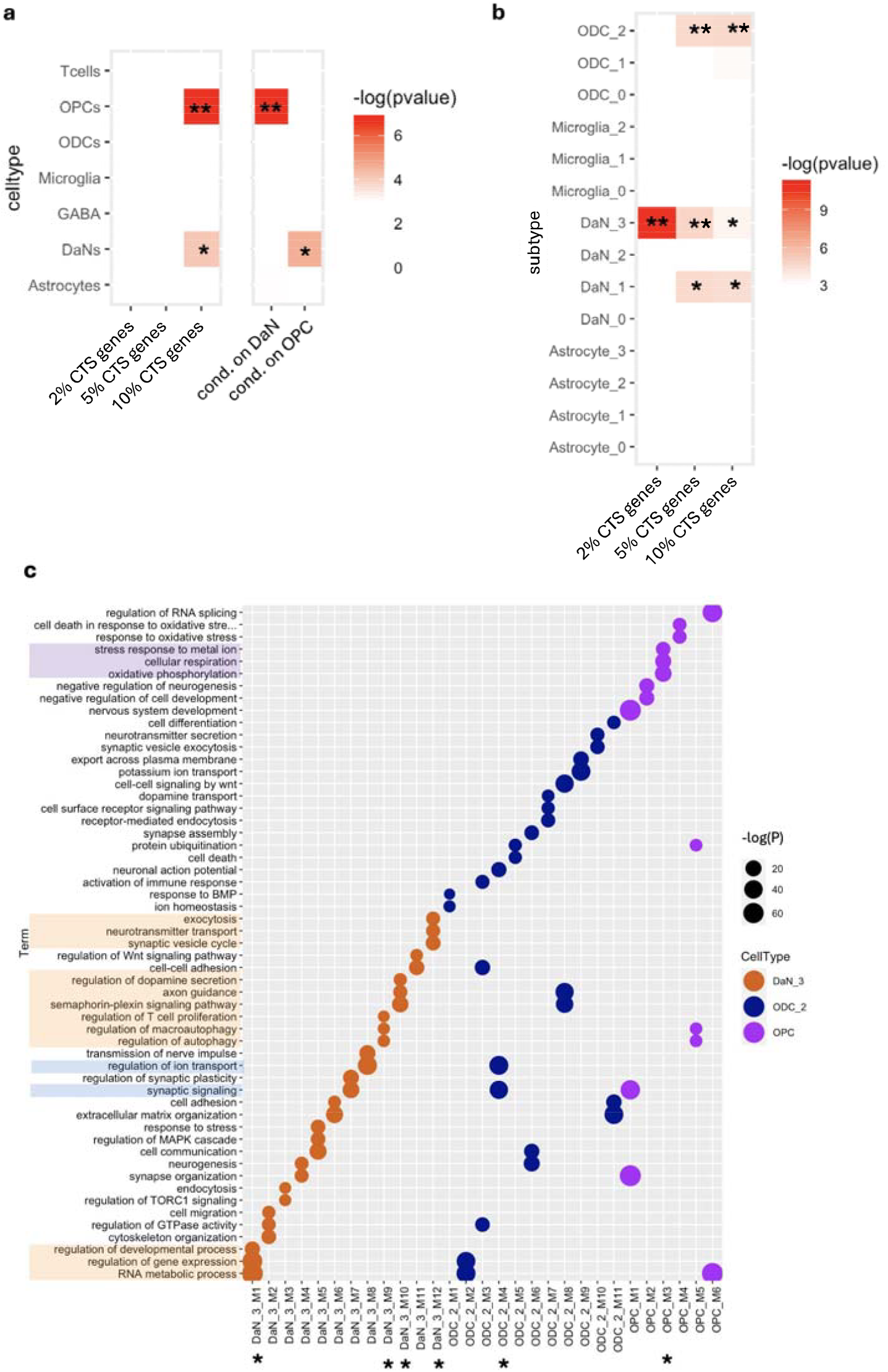
Cell type-specific and pathway-specific PD genetic risk association. **a)** Heatmap showing-log(p-value) obtained by MAGMA for PD genetic risk association with main cell type and **b)** subtype. Double asterisk indicates significance after multiple testing correction at corrected p-value < 0.05, while single asterisk indicates significance at nominal p-value < 0.05. x axis is the X percent of most cell type specific (CTS) genes. **c)** Dotplot showing top enriched Gene Ontology (GO) terms for any cell type-specific protein protein interaction (PPI) gene modules. The size of the dots represents −log(p-value) given by GO enrichment analysis and the color represents the cell type. Single asterisk indicates significance of PD genetic risk MAGMA association at nominal p-value < 0.05.

To further refine the PD-risk associations within each cell type, we used protein-protein interactions (PPIs) between cell-type-specific genes to form PPI modules capturing convergent functionality (Methods). MAGMA gene set analyses of each module found PD risk to be associated with four AGTR1+ DaN_3-specific gene modules (Fig. 2c) highlighting pathways related to neurotransmitter transport, dopamine secretion, cell communication, autophagy and gene expression regulation. MAGMA psODC_2 associations to PD genetic risk involved one module enriched in ion homeostasis and synaptic signalling (Fig. 2c) suggesting dysregulation in neuronal support. MAGMA PD risk significantly converged in one OPC-specific module related to cellular respiration and oxidative process (Fig. 2c) pointing to their higher vulnerability to oxidative damage than other brain cells^33^.

Gene-level expression analysis lacks the sensitivity to detect possible changes at the transcript-level caused, for example, by alterations in alternative splicing^34^. To overcome this limitation, we used differential transcript usage (DTU) analysis to identify additional cell type-specific gDTUs in control samples (Methods). MAGMA PD genetic risk association analysis at transcript-level revealed different sets of risk-associated genes in AGTR1+ DaN_3, psODC_2 and OPCs (Fig. S4) suggesting a complementary signal that can contribute to unravelling gene expression alteration associated to disease. Of note, among the top MAGMA PD risk associated gDTUs in AGTR1+ DaN_3 we found cathepsin B (CTSB) whose blood brain barrier-permeable inhibitors demonstrated efficacy in treatment of many neurological diseases likely given its role in inflammatory response^35^ and in lysosomal pathways^36^.

### Altered cellular landscape of DaNs and ODCs in the ventral SNpc of PD

As with previous midbrain and substantia nigra human atlases^12,13^, we did not detect significantly fewer dopaminergic neurons in PD Braak stage 5-6 samples as compared to control samples (re-analyses of published atlases; Fig. S5). However, many of the cell subtypes associated with PD risk in healthy controls appear affected in the ventral SNpc of PD cases. We first assessed the cell type proportion changes between PD Braak stage 5-6 and healthy control samples for all cell populations (Methods, Table S14). The largest statistically significant reduction was for the AGTR1+ DaN_3 confirming the loss reported^6^, but reductions were also found for the Astrocyte_2 and psODC_2 (Fig. 3a). The spatial location of AGTR1 mRNA was investigated using RNAscope and was found largely in the cell bodies and nuclei of melanised and non-melanised neurons in the SNpc (Fig. 3b). Positive controls show membrane-bound AGTR1 and good preponderance in human kidney peritubular cells (Fig. S1b). AGTR1 transcripts were significantly reduced in PD Braak stage 5-6 compared to healthy controls (p<0.05) (Fig. 3b). AGTR1 (AT1) protein was expressed widely in the neurons but also in the parenchyma (Fig. 3c) and AGTR1 coverage was significantly reduced in PD versus controls and iLBD groups (p<0.05) (Fig. 3c). In healthy controls, perineuronal oligodendrocytes are located within a 5-10 μm distance from the neuronal cell body (Fig. 3d) and on average 1-2 oligodendrocytes are detected per neuron (Fig. 3d). After correction for neuronal loss (Methods), perineuronal oligodendrocytes are significantly decreased in PD versus healthy controls (Fig. 3d). While we are not powered to detect significant differences in cell-type-specific nigral nuclei numbers except between controls and PD Braak stage 5-6, our neuropathological counts reveal that AGTR1-expressing neurons are already significantly reduced in Braak stages 1-4 while psODC_2 are only reduced in PD Braak stage 5-6, showing neuronal loss precedes psODC_2 loss (Fig. 3c,d).

**Figure 3.**
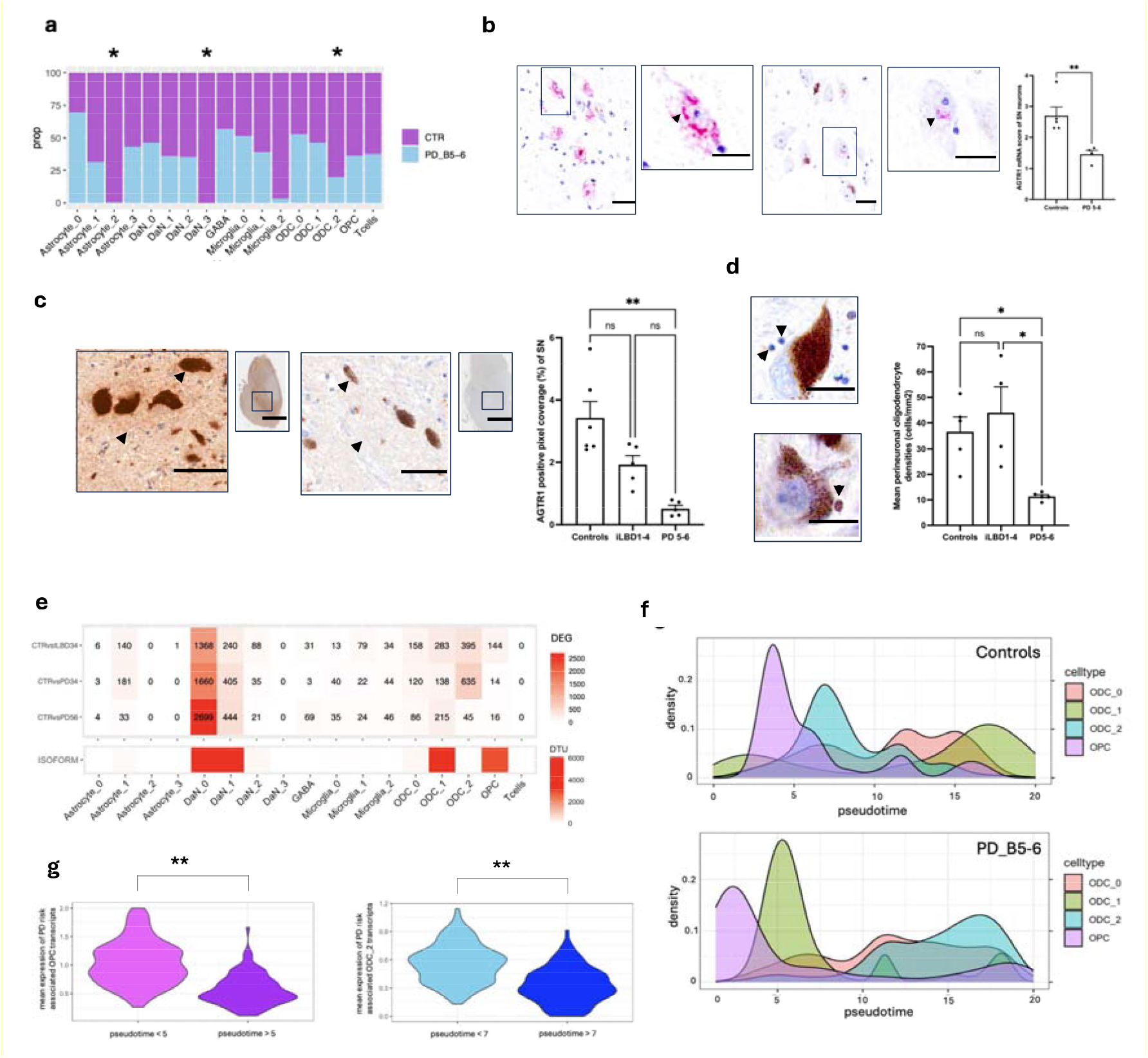
Cell type specific nigral changes in PD. **a)** Barplot showing cell type and subtype proportions in controls and PD_B5-6 samples. Asterisks represent statistically significant proportion changes between the two disease conditions by intersecting significance at FDR-corrected p-value < 0.05 from ScProportion test and at nominal p-value < 0.05 from propeller test (Methods). **b)** Distribution of AGTR1 RNA puncta in the soma of nigral neurons, with a significant loss (n=5 controls; n=4 PD, p<0.01) of AGTR1 puncta in PD versus controls; scalebar=25 µm. **c)** Loss of AGTR1 coverage in the substantia nigra of PD patients (n=5) compared to iLBD (n=5) and healthy controls (n=6) (p<0.01). **d)** Representative image of perineuronal oligodendrocytes surrounding a nigral melanised neuron (top) and immunohistochemistrywith Olig2 (bottom). Perineuronal oligodendrocyte loss in the PD patients (n=4) compared to iLBD (n=4) and healthy controls (n=5), corrected for the neuronal loss in the substantia nigra (p<0.05); scalebar= 25 µm. **e)** Cell type and subtype-specific number of DEGs at abs(FC) > 1 and p value < 0.05 between PD_B5-6 and controls, PD_B3-4 and controls, ILBD_B3-4 and controls (top). Cell type and subtype-specific number of differentially used transcripts (DTUs) at fdr < 0.05 between PD_B5-6 and controls (bottom). **f)** Density plots showing pseudotime trajectory for OPCs differentiation into ODCs computed using transcript expression in controls (top) and PD_B5-6 samples (bottom). **g)** Violin plots showing the mean expression values of OPC-specific transcripts associated to PD risk across two pseudotime intervals in control samples from Fig. 3f (left) and the mean expression values of ODC_2-specific transcripts associated to PD risk across two pseudotime intervals in control samples from Fig. 3f (right). PD risk-associated transcripts are up-regulated in the early stages of OPC differentiation and in ODC_2 maturation. Asterisks indicate Wilcoxon test p-value < 2.2e-16.

We next determined differentially expressed genes (DEGs) between PD patients and healthy controls in each cell population (with the exception of AGTR1+ DaN_3 and Astrocyte_2 due to their complete loss/absence in the disease samples) (Fig. 3e). OPCs, Microglia, GABAergic neurons, T-cells and Astrocytes (except for Astrocyte_1) showed little disease effect at gene expression level. In contrast, DaN_0 and DaN_1 showed the highest number of DEGs across all disease stages. DaN_2 exhibited few DEGs appearing relatively disease-resistant concordant with a protective role associated with the TMEFF2 DaN_2 marker gene^37^. ODCs followed with a relatively high number of DEGs, although psODC_2 only showed dysregulated gene expression at iLBD Braak stage 3-4 and PD Braak stage 3-4, noting that the reduction in psODC_2 in PD Braak stage 5-6 likely affects power to detect DEGs. Functional enrichment analysis revealed an overall downregulation of cell-cell signalling, cell communication and neuronal support, and upregulation of mitochondrial-related processes specifically in the DaN_1 population (Table S4). This agrees with trajectory analysis of DaN_0 and DaN_1 showing downregulation of synaptic signalling and upregulation of oxidative phosphorylation along disease progression (Fig. S6, Table S6, Suppl. Note). Astrocyte_1 showed a specific enrichment in response to cytokine, ODC_0 in response to unfolded protein, cell activation and regulation of cell death, and ODC_1 in stress response signalling (Table S4).

Analyses of differential transcript usage (DTU) in the ventral SNpc in PD as compared to healthy controls revealed cell type specific alterations distinct from DEGs (Fig. 3e, Methods). We observed a significantly higher number of genes with differential transcript usage (gDTU) than DEGs in OPCs and ODC_1 (Fig. 3e) with only 1% in OPCs and 13% in ODC_1 overlapping gDTU and DEGs, respectively (Fig. S7). DaN_0 and DaN_1 also showed a high number of gDTUs but a greater overlap with DEGs with 24% and 26% overlapping gDTUs and DEGs, respectively (Fig. 3e, Fig. S7). We noted a high degree of shared gDTUs across different cell populations (Fig. S8) mainly enriched in cellular response to stress, cell morphogenesis, regulation of MAPK cascade, vesicle-mediated transport and RNA splicing (Table S4). Relevant to downstream analyses, OPCs showed strong unique enrichment in cell developmental processes (Table S4) with the top gDTU being anosmin-1 (ANOS1) whose over-expression is reported to regulate OPC proliferation and migration^39^.

### Differential isoform usage affecting genes associated with OPC-ODC differentiation provides a mechanism for loss of psODC_2 in PD

Given the significant differential gene isoform usage in OPCs and ODCs (Fig 3e), we performed isoform-based pseudotime trajectory analysis across OPC and ODC cell types separately for PD Braak stages 5-6 and healthy controls (Methods). We confirmed the position of the marker genes for OPCs (PDGFRA and PCDH15), for committed OPCs (PTPRZ1 and VCAN) and for newly formed ODCs (BMP4 and SCN2A) along the differentiation trajectory (Fig. S9). We observed altered ordering in PD, with psODC_2 adjacent to OPCs in healthy controls but ODC_1 adjacent to OPCs in PD suggesting altered OPC-ODC differentiation (Fig. 3f). Supporting this, OPC-specific gDTUs between PD Braak stages 5-6 and healthy controls were enriched in biological processes such as cell development, regulation of cell cycle and lipoprotein transport (Table S4). Furthermore, OPC-specific MAGMA PD genetic risk-associated transcripts (Methods) were upregulated early along the OPC trajectory that correlated with differentiation and were enriched in the Smoothed signalling pathway, stem cell population maintenance and developmental processes (Fig. 3g, Table S5). psODC_2-specific MAGMA PD genetic risk-associated transcripts (Methods) were instead upregulated at the point of transition from OPCs to psODC_2 and enriched in inhibition of maintenance of pluripotency, apoptotic processes involved in development and response to hypoxia (Fig. 3g, Table S5) supporting dysregulation of both OPC differentiation and psODC_2 maturation as contributing factors to psODC_2 reduction in late-stage PD samples.

### Perineuronal satellite psODC_2 and OPCs provide a supportive environment to AGTR1+ DaN_3 that is disrupted in PD and associated with genetic risk

Given the strong enrichment in cell-cell signalling pathways across different cell populations affected in PD, we next sought to predict cell-cell communication networks by identifying ligand/receptor (LR) interactions between source and target cells in healthy controls and in PD Braak stage 5-6 samples (Methods). Within healthy control tissue, we found significant enrichments for MAGMA PD genetic risk-associated genes in LR interactions between AGTR1+ DaN_3, OPC and psODC_2, between psODC_2 and Astrocyte_1, and between OPC, psODC_2, Microglia_2 and Astrocyte_1 (Fig. 4a,b, Methods). The crosstalk between AGTR1+ DaN_3 and psODC_2 mainly involved laminin, plexin, EPHA and CDH signalling pathways (Fig. 4b,f) that promote neuronal survival^40,41^ with their ligands and receptors being downregulated early along the psODC_2 disease-specific trajectory (Fig 4c, Suppl. Note, Fig. S10, Methods). Similarly, the crosstalk between AGTR1+ DaN_3 and OPCs mainly involved OPC receptors involved in maintenance and survival of neurons (Fig 4b,f). When comparing the differential number and strength of LR interactions between PD and controls, we observe global decrease of cell communications in PD mainly caused by the loss of AGTR1+ DaN_3 as the core cell population in the identified networks (Fig. 4d,e). Likely relevant to their loss, most of the AGTR1+ DaN_3-specific incoming signalling pathways were enriched in neuronal development, survival and neuroprotection (CDH, EPHA, EPHB, APJ/APELIN, HSPG, APRIL, VEGF, Semaphorin, ALCAM, Wnt, HSPG), in synaptic plasticity (Reelin) and dopamine release (CXCL) (Fig. 4f, Fig. S11). Similarly, active crosstalk between AGTR1+ DaN_3 and Astrocyte_1 involved AGT, VEGF, SEMA4, neurothrophic factors and APRIL signalling pathways (Fig. 4f, Fig. S11) characteristic of astrocyte reactivity sustaining neuronal survival under stress conditions^42,43^.

**Figure 4.**
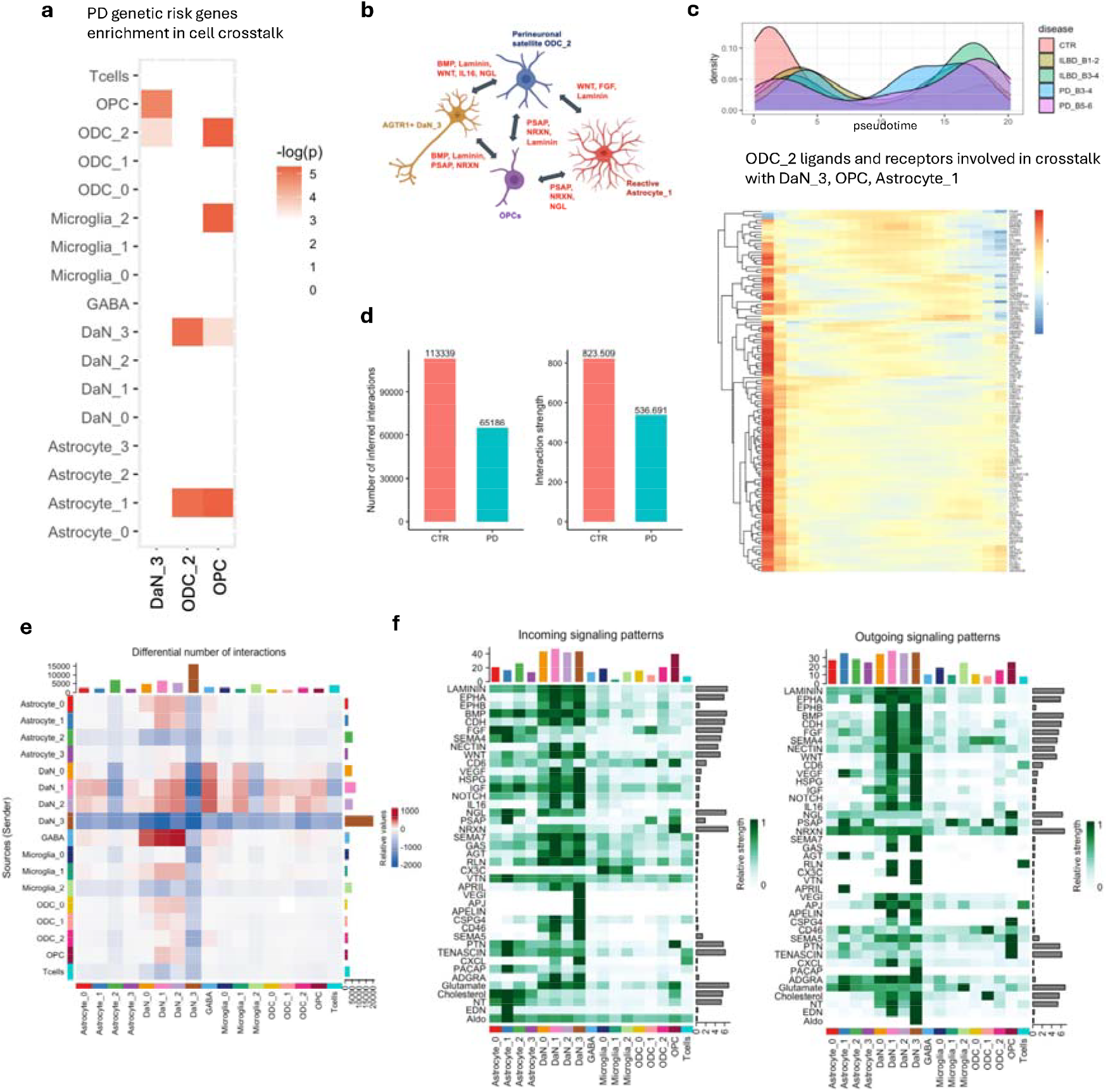
Cell crosstalk between psODC_2, OPCs and AGTR1 DaNs is affected in PD. **a)** Heatmap showing-log(p-value) for the enrichment in DaN_3-, ODC_2- and OPC-specific MAGMA PD risk associated genes of cellChat derived ligand/receptor interactions between DaN_3, ODC_2, OPCs and any other cell type. **b)** Network of signaling pathways affected by PD genetic risk. **c)** Heatmap showing significant expression changes along ODC_2 disease progression trajectory (top, Suppl. Note, Fig. SB) of ODC_2-specific ligands and receptors involved in crosstalk with DaN_3, OPC and Astrocyte_1. **d)** Number of inferred interactions (left) and interaction strength (right) betweenPD_B5-6 samples and controls. **e)** Heatmap showing differential number of interactions between PD_B5-6 and controls between any cell type and subtype. **f)** Heatmaps showing the summary of selected signalling pathways that contribute to outgoing (right) or incoming (left) communication. The colour bar represents the relative signalling strength of a signalling pathway across cell types. The bars indicate the sum of the signalling strength of each cell type or pathway.

### A metabolic stress state underlies DaN-specific vulnerability

Among the four DaN subtypes, AGTR1+ DaN_3 showed the highest expression of genes involved in dopamine signalling (Fig. 5a) and of the main effectors of the renin-angiotensin system (RAS) (Fig. 5b). Given the increased neuronal support signalling, we hypothesised that AGTR1 expression indicates a stressed cell *state*, rather than *type*. To test this hypothesis, we challenged iPSC-derived dopaminergic neurons in vitro with PD-relevant stressors, MPP+ and 6-OHDA, and indeed detected a significant increase in AGTR1 protein (Fig. 5c, Methods). Given the high gene expression similarity and close proximity in UMAP plot of DaN_1 to DaN_3 populations (Fig. S12 and Fig. 1d), we hypothesised a transitioning process from the DaN_1 population towards a highly metabolic active AGTR1+ DaN_3 cell state that is vulnerable in PD. In fact, by examining DaN_1/DaN_3 pseudotime trajectory in healthy control samples (Methods) we found a large overlap between the two DaN populations but also higher proportion of DaN_1 towards lower pseudotime values and of DaN_3 towards higher pseudotime values proposing that DaN_1 neurons can transform into more synaptically active DaN_3 neurons (Fig. 5d). We noted that not only AGTR1 but most of the RAS-related genes were upregulated along this trajectory (Fig. S13). Furthermore, functional analysis of gene modules that correlated with the identified trajectory highlighted upregulation of synaptic signalling, plasticity and transmission, dopamine transport, endocytosis, cell communication, neuronal development and migration, but also stress-activated MAPK cascade, JNK cascade, immune response, gliogenesis and macroautophagy at the point of cell transition and later (Fig 5e, Table S5, Table S6). Notably, these processes are also enriched in MAGMA PD genetic risk genes (p-value<0.05, Fig 5e, Methods) relatively early along the cell-state transition trajectory. As AGTR1+ DaN_3 neurons account for a small percentage of ventral SNpc DaNs in healthy controls, their loss could not account for the estimated >50% of DaNs lost in PD patients upon diagnosis^5^. However, as a vulnerable DaN state, rather than type, we propose that nigral DaNs transition to this state prior to loss. This hypothesis may also explain the efficacy of ARBs treatment in reducing PD risk in patients with ischemic heart disease and high blood pressure as recently reported^44,45,46^; ARBs may act to block the transition to this vulnerable state thereby reducing DaNs loss.

**Figure 5.**
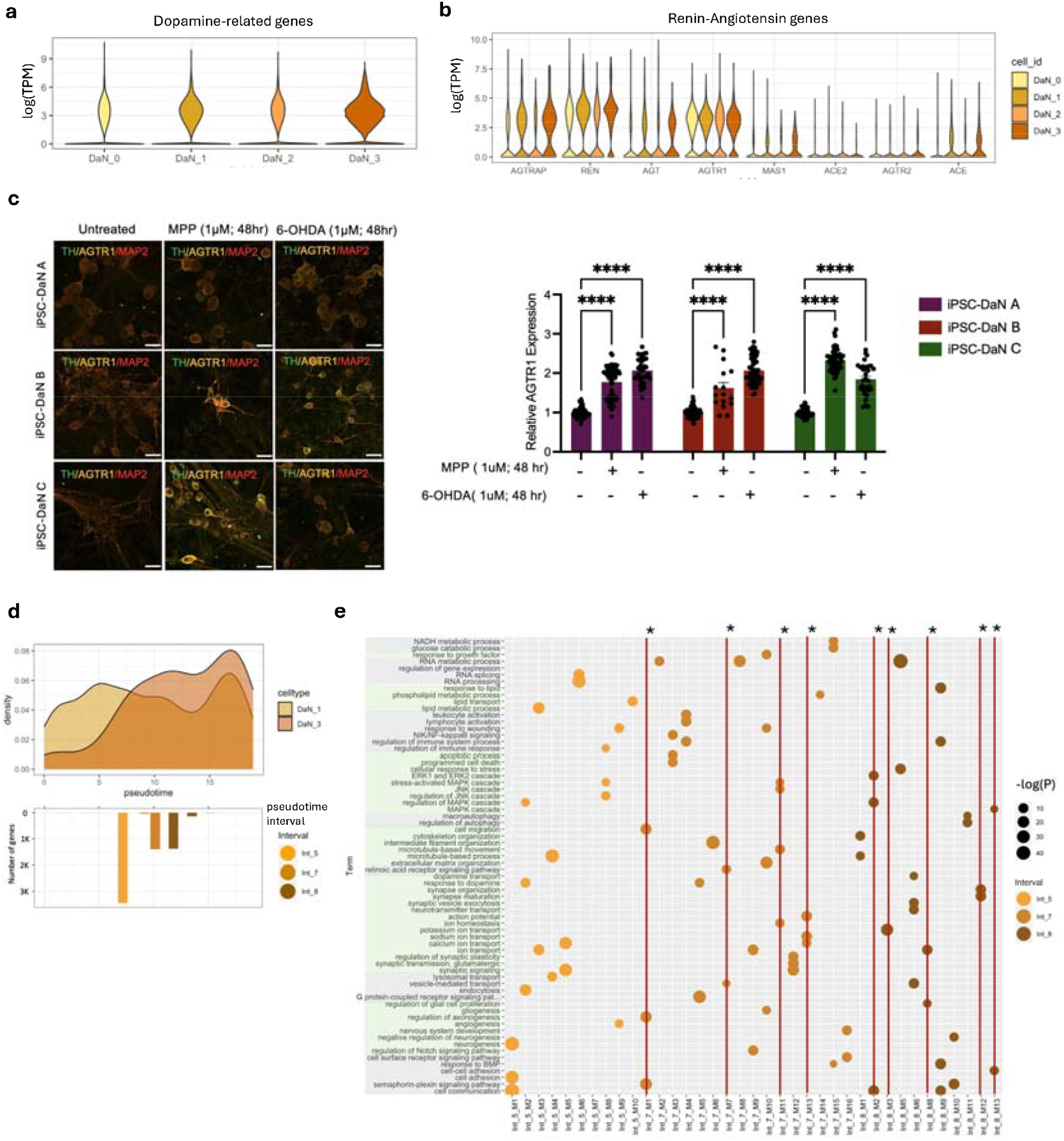
Cell state transition toward highly vulnerable AGTR1 DaN_3. **a)** Violin plot showing the expression of dopamine pathway-related genes by DaN subtype. **b)** Violin plot showing the expression of main renin-angiotensin pathway-related genes by DaN subtype. **c)** Representative confocal images of iPSC-derived DaNs (three lines denoted as A, Band C) treated with PD inducers MPP+ Iodide or 6-OHDA and stained for Tyrosine Hydroxylase (TH), pan neural marker MAP2 and AGTR1 (left) and subsequently quantified for AGTR1 expression levels in the DaNs (right). Data represented as mean± SEM. **d)** Density plot showing pseudotime trajectory for AGTR1+ cell state transition from DaN_1 subtype (top). Barplot showing the number of statistically significant genes changing expression across intervals identified along the pseudotime trajectory (bottom). **e)** Dotplot showing enriched GO terms for protein protein interaction (PPI) modules built on genes changing expression across intervals identified along the pseudotime trajectory. The size of the dots represents −log(p-value) for GO enrichment and the color represents the three main intervals on the trajectory. Single asterisk indicates MAGMA PD risk significant association at nominal p-value < 0.05 for the protein protein interaction (PPI) modules.

### AGTR1+ DaN_3 and perineuronal ODCs at the intersection of PD and insulin resistance

Activation of RAS involves stimulating kinase pathways, such as mitogen-activated protein kinases (MAPK, p38) and JAK/STAT pathways^47^ which have a crucial role also in diabetes and diabetes-related cardiovascular complications^48^. Beyond PD risk, we also identified convergent enrichment of metabolic genetic risk in both AGTR1+ DaN_3 and perineuronal ODC_2 populations, with the strongest association with triglyceride levels (TG), a proxy for insulin resistance (Fig. 6a). Conditional analyses for AGTR1+ DaN_3 revealed that, PD, T2D, HbA1c (hyperglycaemia) and TG risk loci were associated with largely distinct gene sets. By contrast, in perineuronal ODC_2, TG-associated loci appeared to account for much of the observed PD genetic overlap, removing or reducing the associations with PD and other metabolic traits and demonstrating a greater degree of shared risk-associated genes in this cell type (Fig. 6b). In perineuronal ODC_2, PD and metabolic risk converged on retinoic acid and ephrin receptor signalling, regulation of synaptic plasticity and neurotransmitter secretion, glutamate receptor signalling, Hippo pathway regulation, and broader cell–cell communication processes (Fig. S14). Notably, many of these shared pathways have established crosstalk with the PI3K/AKT/mTOR axis, a signalling hub regulated by insulin, IGF, BDNF, NGF, and glutamate, with many members significantly downregulated in perineuronal ODC_2 in PD brains as compared to healthy controls and proposing an insulin resistant state (Fig. 6c)^49^. Consistent with this, we also observed significantly higher expression of the insulin receptor, INSR, in perineuronal ODC_2 as compared to other nigral cell types, highlighting insulin sensitivity (Fig. 6d). Given the central role of PI3K/AKT/mTOR signalling in promoting cell survival and regulating responses to metabolic and oxidative stress^50^, these findings propose that dysfunction of this pathway particularly in perineuronal ODC_2 may underlie the heightened metabolic vulnerability of both AGTR1+ neurons and perineuronal ODCs in Parkinson’s disease.

**Figure 6.**
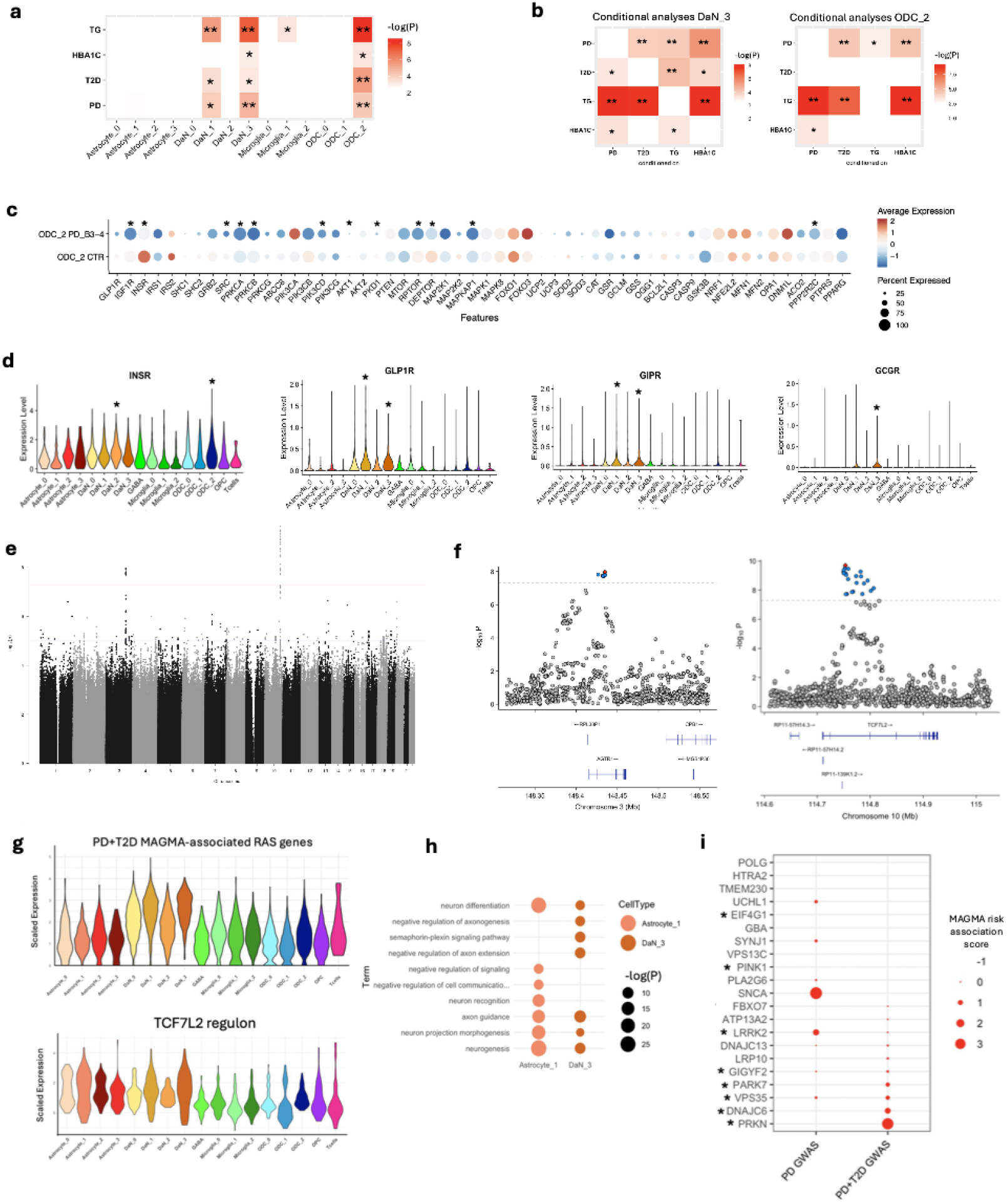
AGTR1+ DaN_3 and perineuronal ODC_2 states at the intersection of PD and metabolic traits. **(a)** Heatmap showing −log(p-value) obtained by MAGMA for PD,T2D, HBA1C andTG genetic risk association with cell subtype. Double asterisk indicates significance after multiple testing correction at corrected p-value < 0.05, while single asterisk indicates significance at nom inal p-value <0.05. **(b)** Heatmap showing −log(p-value) obtained by MAGMA for pairwise conditional analyses between PD, T2D, HBA1C andTG genetic risk association with DaN_3 and ODC_2. Double asterisk indicates significance after multiple testing correction at corrected p-value < 0.05, while single asterisk indicates significance at nominalp-value < 0.05. **(c)** Dotplot showing the expression of key receptors and downstream effectors of the PI3K/AKT/MTOR pathway in perineuronal ODC_2 across PD_B3-4 cases and healthy controls. Single asterisk indicates significance at FDR-corrected p-value < 0.05 for differentially expressed genes between PD_B3-4 cases and healthy controls, **(d)** Violin plot showing the expression of insulin receptor and of T2D GLP1 agonists targets by cell type. Single asterisk indicates significance at FDR-corrected p-value < 0.05 for differentially expressed genes, **(e)** Manhattan plot of the meta-analysis results between Parkinson patients with a history of type 2 diabetes vs no history of type 2 diabetes in three different population cohorts. The x axis shows chromosome and base pair positions of each variant tested in the meta-analyses. The y axis shows the P value in the −log10 scale. Red horizontal line indicates the genome-wide significant threshold of P < 5 × 10-8 and the blue line indicates the suggestive threshold of 1 × 10-5. **(f)** Region plots depicting genome-wide significant loci on Chromosome 3 (left) and Chromosome 10 (right), (g) Violin plot showing the expression of PD/T2D MAGMA-associated RAS genes (top) and ofTCF7L2 regulon by cell types (bottom), **(h)** Dotplot showing top enriched GO terms in targets genes predicted to be regulated by TCF7L2 in AGTR1+ DaN_3 and Astrocyte_l. **(i)** Dotplot showing known PD rare variant genes according to their association with sporadic PD GWAS and with comorbid PD and T2D GWAS. The size of the dots represents the scaled MAGMA association score (ZSTAT). Asterisk indicates that a gene is associated to non-Lewy body pathology.

Recently, GLP1R agonists used to treat T2D have been proposed as candidate treatment for PD patients^51^. We found GLP1R upregulated along with other T2D therapeutic targets like GCGR and GIPR specifically in the AGTR1+ DaN_3 population (Fig. 6d). Individuals with T2D have a significantly increased risk of PD^52^. Given the T2D genetic risk MAGMA association with AGTR1+ DaN_3 specific gene expression, we examined the risk associated with PD comorbid with T2D. We performed a whole genome wide association study (GWAS) between PD patients with a history of T2D vs PD patients with no history of T2D combining three different PD cohorts: the Oxford Parkinson Discovery Cohort^53^, including 965 PD with 69 with a history of diabetes; the Tracking cohort^54^, including 1436 PD with 97 with history of diabetes; and 5460 participants of the UK Biobank (UK BB) who received a diagnosis of PD since 2023 including 866 with a history of diabetes (Methods). The QQ plot showed a slight deviation from the diagonal line across the entire range, but a strong deviation at the tail, which could suggest potential inflation of p-values due to true signals or systematic bias (Fig. S15). The GWAS identified two genome-wide significant associations, in the AGTR1 and the TCF7L2 loci (Fig. 6e,f). MAGMA associated PD+T2D risk genes were enriched in the RAS pathway (p-value=1.2 × 10^-5^) with these RAS genes showing higher expression in AGTR1+ DaN_3 than other cell types (Fig. 6g). The direct association of AGTR1 and other RAS genes with comorbid PD and T2D, the association of T2D genetic risk with DaN_3 gene expression profiles and the upregulation of T2D-associated molecular processes in DaN_3 provides compelling evidence for a genetic and molecular association between PD and T2D through this vulnerable cell state. TCF7L2 is a transcription factor (TF) and a long known T2D risk gene that has been described not only to have a role in development and survival of DaNs^55,56,57^ but also to regulate the transcription of the gene encoding the insulinotropic hormone GLP-1 in vitro^58^ acting as ligand to the T2D therapeutic target GLP1R. TCF7L2 is highly expressed in astrocytes, particularly in reactive Astrocyte_1, and among DaNs its expression was highest in DaN_3 (Fig. S16). We used single-cell regulatory network inference (SCENIC) to identify highly specific regulons to the DaN and Astrocyte subtypes. SCENIC revealed that TFC7L2 was among the top most active TFs ranked by AUC specifically in the AGTR1+ DaN_3 and in Astrocyte_1 (Fig.6g, Fig. S17, Table S9, Methods) with target genes enriched in neuron differentiation, axon guidance and semaphorin-plexin signalling pathway in AGTR1+ DaN_3 (Fig. 6h, Table S9), and in neuron recognition, communication and differentiation in Astrocyte_1 (Fig. 6h, Table S9). Functional analysis of the top MAGMA associated genes to this fraction of T2D genetic risk revealed significant enrichments in DaN_3 specific cellular pathways related to neuron development, synaptic transmission and regulation of Hippo signalling (Fig. S18, Table S10, Methods), which plays a role in function and survival of dopaminergic neurons and their response to oxidative stress^59^. Finally, we examined known familial PD genes for their association with comorbid PD and T2D genetic risk using MAGMA. Many known familial PD genes were associated with comorbid PD and T2D genetic risk, namely *PRKN, DNAJC6, PARK7* and *VPS35* which are involved in dopamine metabolism and stress response (Fig. 6i, Methods). Notably, comorbid PD and T2D genetic risk associated familial PD genes give rise to non-Lewy body pathology^60,61^ Parkinsonisms, proposing a distinct synuclein-independent aetiology.

## Discussion

Parkinson’s disease is a complex neurodegenerative disorder characterised by an interplay of genetic, metabolic, and cellular factors contributing to neuronal loss in SNpc. While recent work has identified AGTR1+ dopaminergic nigral neurons as selectively vulnerable in PD^6^, the mechanisms underlying this susceptibility have remained elusive. Here, through deep single-nuclei sequencing of the ventral SNpc across PD stages, combined with novel GWAS data, we uncover a network of genetic and cellular interactions that underly this susceptibility. Our findings reveal that PD genetic risk converges in AGTR1+ neurons, in perineuronal satellite oligodendrocytes (psODCs)—a little-studied, non-myelinating oligodendrocyte population— that are both lost in PD, in oligodendrocyte precursor cells (OPCs) and, to some degree, in reactive astrocytes (Fig. 2a,b, Fig. 4a). Importantly, we also validate our findings histologically and show a significant loss of AGTR1 transcripts and protein, as well as a loss of psODCs in the SNpc in PD patients compared to healthy controls (Fig. 3b,c,d). PD genetic risk also converges in potentially targetable crosstalk between these same cell subpopulations supporting a broader disruption of neuron-glia interactions (Fig. 4a). Furthermore, pseudotemporal ordering reveals altered OPC to ODC differentiation with widespread changes in isoform usage in genes underlying OPC/ODC differentiation pathways within PD which may underlie the observed reduction in psODCs (Fig. 3f). These transcriptomic changes in glial cells represent a novel dimension of PD pathology, suggesting that oligodendrocytes play a more active role in dopaminergic neuron survival than previously appreciated.

The original associations of PD common genetic risk with the expression profiles of ODCs and OPCs in both early human and mouse cellular brain atlases was unexpected and unexplained^10,62^. However, it may have been unappreciated that psODCs are thought to outnumber astrocytes amongst cells in close proximity to neurons and that OPCs are reported specifically to be in close proximity to nigral dopaminergic neurons^38,63^. We reveal a tight coupling of these proximal cell types’ intercellular communication. For OPCs, AGTR1+ DaN_3 were found to engage in crosstalk particularly through OPC-expressed receptors implicated in neuronal maintenance and survival (Fig. 4b,f) and with the OPC-expressed ligand CSPG4 influencing cell adhesion and survival through integrin binding on AGTR1+ DaN_3 (Fig. 4f). This observation aligns with emerging evidence that OPCs may exert neuroprotective effects independent of their differentiation into mature oligodendrocytes^63^. Recent studies have demonstrated that OPCs can form specialised contacts on neuronal somata that facilitate the exocytosis of neuronal lysosomes, thereby contributing to neuronal homeostasis and potentially offering a novel mechanism of neuroprotection in neurodegenerative contexts^64^. Similarly, crosstalk between AGTR1+ DaN_3 and psODCs is predominantly mediated by signalling pathways involving laminin, plexin, EPHA, and cadherins (CDH), all of which are known to play crucial roles in supporting neuronal survival (Fig. 4b,f)^40,41^. Some of these neuroprotective pathways were both enriched in PD genetic risk variants and progressively downregulated from early disease stages along the psODCs disease-associated trajectory (Fig. 4c, Suppl. Note, Fig. S10, Methods). The overlap between PD genetic risk and triglyceride level trait genetics (TG; Fig. 6a,b) identifies shared risk between PD and insulin resistance focussed within psODCs, which is pathologically supported by lowered PI3K/AKT/mTOR pathway gene expression during disease providing an explanation for the role of ODCs in PD.

A key proposal of our study is that AGTR1+ neurons should be defined as a cell state rather than a distinct cell type, characterised by elevated dopamine synthesis and increased metabolic stress responses. Specifically, these neurons exhibit activation of the renin-angiotensin system and the MAPK cascade, both of which contribute to oxidative stress and mitochondrial dysfunction, contributing to selective vulnerability in PD ^65,66^ (Fig. 5). Our genetic analyses establish a direct link between T2D and PD, with genome-wide significant loci within AGTR1 and TCF7L2 (Fig. 6). We provide a mechanistic rationale for the observed significant neuroprotective effects of blood-brain barrier-crossing angiotensin receptor blockers (ARBs)^67,68^ and GLP-1R agonists in PD^21^. Both GLP-1R agonists and ARBs have been shown to rescue dysfunction in PD models^22,69^. Indeed, we show upregulation of both GLP1R and GIPR in vulnerable AGTR1+ DaN_3 neurons (Fig. 6d) suggesting dual GIP/GLP-1 receptor agonists may be most efficacious^70^. More immediately, as RAS-targeting drugs are amongst the most commonly prescribed (8-35% of adult populations)^71^, typically prescribed in the 40-60 age range prior to the average age of PD diagnosis of about 70^72^, a change in the clinical practice of treating hypertension away from ACE inhibitors and non-blood brain barrier (BBB) penetrant ARBs towards BBB-penetrant ARBs could significantly reduce the risk of PD among a large fraction of the population without any preselection for PD risk required.

Although progress has been made over recent years in identifying disease-modifying therapies for PD, targeting particularly α-synuclein, these approaches have yet to meet clinical endpoints^73^, likely due to the heterogeneity of PD with diverse genetic susceptibilities, risk factors, multiple pathogenic mechanisms and clinical trajectories. Notably, whether α-synuclein aggregation is a key feature for the development and progression of the disease remains under debate. Not all cases of parkinsonism feature Lewy pathology (e.g., PRKN- and half of the LRRK2-mutation carriers), yet neurodegeneration still occurs^60,61,74^. Furthermore, Lewy bodies and neurites can be present in individuals without clinical PD symptoms^75^, raising questions about their causal role in neurodegeneration. Our findings support an alternative paradigm where a significant component of PD is driven by metabolic stress and mitochondrial dysfunction, rather than α-synuclein pathology associated with the presence of Lewy bodies. Relevant here, we invoked AGTR1 expression in iPSC dopaminergic neurons using the toxin MPTP/MPP+ that causes a non-Lewy body Parkinsonism^76^. Our paradigm aligns with the known PD familial risk genes that were significantly associated with comorbid PD/T2D risk —such as PRKN, PARK7, DNAJC6 and VPS35—which predominantly affect mitochondrial function, oxidative stress responses and dopamine homeostasis and are not associated with Lewy pathology^60,61^. Of the remaining familial PD genes showing some association with non-Lewy pathology and not associated with either the common genetic risk of Parkinson’s or comorbid PD/T2D risk (Fig. 6i), PINK1 physically interacts with PRKN, while EIF4G1 regulates glucose homeostasis^77^. This paradigm proposes a synuclein-independent metabolic vulnerability but does not exclude exacerbating synucleinopathic interactions; given the association of sporadic PD genetic risk with AGTR1+ dopaminergic neurons and their reported wide-spread loss in PD patient nigra^6^ these interactions may make a significant contribution to PD aetiology.

Given the metabolic underpinnings of PD vulnerability, stratification of patients based on neuronal metabolic stress could refine therapeutic strategies. For instance, in vitro data indicate that exenatide (a GLP-1R agonist) suppresses MAPK pathway activation, suggesting that PD patients with high baseline MAPK activation and insulin resistance may benefit most from GLP-1R agonist therapy^22^. Our study offers a significant shift in understanding PD vulnerability, identifying metabolic stress-driven mechanisms that appears distinct from α-synuclein-associated pathology. The interplay between AGTR1+ neurons, psODCs, astrocytes, and diabetes-associated metabolic pathways suggests that targeting renin-angiotensin signalling, MAPK activation, and insulin resistance, perhaps combinatorially, could open new therapeutic avenues. These insights not only deepen our understanding of PD pathogenesis but also pave the way for precision medicine approaches in neurodegeneration.

## Methods

### Human brain tissue samples

Post-mortem brain tissues from patients with PD, iLBD, and healthy controls were obtained from the Oxford Brain Bank (OBB, University of Oxford, UK) and the Parkinson’s UK Brain Bank (Imperial College London, UK), in accordance with approved protocols by the South Central - Oxford C Research Ethics Committee (ref 23/SC/0241) and London Multicentre Research Ethics Committee, respectively. All participants provided informed consent for the brain donation. Both brain banks comply with the requirements of the Human Tissue Act 2004 and the Codes of Practice set by the Human Tissue Authority (HTA licence numbers 12217 for OBB and 12275 for the Imperial). The clinico-pathological demographics of the patients and healthy controls are summarized in Table S1. The tissue sampling method is detailed in Fig. S1a, which shows the careful procedure obtaining dopaminergic neurons from the ventral substantia nigra pars compacta (SNpc) for sequencing.

### Cell lines and ethics committee approval

The human iPSC (hiPSC) lines utilised in the study are detailed in Table S11. These lines were acquired from the European Stem Cell Biobank (EBiSC). Human iPSC lines were cultured under feeder-free conditions on Geltrex-coated 6-well plates, using mTeSR Plus medium and passaged every 4-5 days with 0.5 mM EDTA solution or upon reaching 80% confluency. A passage number between 20 and 30 was employed for differentiation purposes.

### Nuclei extraction

Nuclei were isolated from frozen human post-mortem brain punches by tissue homogenisation on ice in 400 µl of homogenisation buffer using a pre-chilled dounce homogenizer and pestle by gentle strokes until dissolved. The homogenate was filtered to remove debris greater than 30 µm in size and centrifuged for 5 min 400 g at 4°C. The supernatant was discarded and the pellet resuspended in 500 µl of staining buffer. 200 µl was then removed to use as an unstained control in FACS, and the remaining 300 µl was stained with 1 µl Hoechst stain. Nuclei were isolated by flow cytometery and resuspended in staining buffer/Nuclei Storage buffer (Sigma) for use in downstream applications.

### snRNA-seq preparation

Cells were dispensed at a concentration of 30,000 cells/mL onto nanowells using the ICELL8 Single-Cell System (Takara). The cDNA libraries were generated using the SMART-Seq ICELL8 cx Application Kit protocol provided by the manufacturer. Sequencing was carried out on Illumina NovaSeq 6000 platform using 100 bp paired-end sequencing with dual indexing.

### snRNA-seq data processing

Raw sequencing data were processed using the Mappa pipeline v1.0 (Takara) and aligned to Homo sapiens GRCh38.99 primary assembly. Briefly, Mappa pipeline consisted in running *demux* for demultiplexing and barcode assignment followed by *analyzer* to obtain gene counts including introns (*Cutadapt* for adapter removal and trimming, *STAR* for genome alignment, *Subread* for counting).

### snRNA-seq data filtering

We noted that on average more than 35% of all reads mapped to introns. These intronic reads were informative about expression levels and significantly increased the number of detected genes (Fig. S19 top) but also generated a baseline expression signal that obscured cluster resolution (Fig. S19 bottom). To account for the background noise and to keep only the most informative signal, we generated two different datasets: one made of all protein coding genes derived from mapping exonic regions only and one made of the top 10000 most variable protein coding genes derived from mapping intronic regions only. We then merged the two gene counts matrices by summing exonic-derived counts and intronic-derived counts for each gene. By doing so, we obtained better defined cell type specific clusters that we could not identified when using the initial exon+intron-based gene counts dataset.

We then removed cells with particularly low number of detected genes (< 2000), and cells with extremely high percentage of the library mapping to mitochondrial genes (> 20%). Also, we removed lowly expressed genes keeping only those with more than 1 count in at least 5% of filtered cells.

### Dimensionality reduction and clustering

We applied Seurat workflow for the integration of scRNA-seq datasets using reciprocal PCA (RPCA)^78^. Firstly, the dataset was split into a list of five seurat objects (one per disease stage). Normalisation and identification of the top 2000 most variable features were done independently for each Seurat object. Anchors between any two datasets were then determined by projecting each dataset into the others PCA space and constraining the anchors by the same mutual neighborhood requirement using FindIntegrationAnchors(). The anchors were used to integrate all datasets together with IntegrateData(). The standard workflow for visualization and clustering was performed on the new integrated dataset. Clusters were annotated to one of seven major cell types (astrocytes, dopaminergic neurons, GABAergic neurons, oligodendrocytes, oligodendrocyte precursor cells (OPCs), T-cells and microglia/macrophages) based on per-cluster expression of a list of known marker genes identified in published datasets. Once major cell types were extracted, subtypes were defined by re-analysing each major cell type separately and applying the workflow above. Clusters were then annotated to cell subtypes based on known marker genes.

### Differential Expression Analysis

To identify cluster markers and differentially expressed genes (DEGs) between experimental groups we used a Wilcox test implemented in the *FindMarkers* function from Seurat R package. DEGs were defined as those with an adjusted p value < 0.05 and abs(logFoldChange) > 1. For cluster markers, where cells within a cluster are compared to the rest, we only considered positive markers.

### Gene Ontology enrichment analysis

We used *TopGO*^*79*^ for Gene Ontology (GO) enrichment analysis, using the respective background gene population of each filtered dataset. The R package rrvgo was used to reduce and visualize lists of GO terms by identifying redundance based on semantic similarity^80^.

### Cell type-specific gene sets

To define cell-type-specific gene sets from the TPM expression matrix we modified the approach of^81^, whereby, for each gene the non-parametric *Mann-Whitney U test* is calculated between the expression of that gene in a given cell population as compared to its expression in all other cells. We then defined cell-type-specific gene sets within the range 2% - 10% of genes with the highest statistic in that cell-type. Note that genes are not necessarily exclusive to a single cell-type.

### Cell type association analysis

We used MAGMA (version 1.09b) to perform gene-set enrichment analysis based on GWAS summary statistics while accounting for LD structure between SNPs^82^. We downloaded publicly available summary statistics for PD^83^ and for T2D from [https://www.diagram-consortium.org/downloads.html]. Specifically, a competitive gene-set analysis linear regression model was performed to test the hypothesis that a cell-type-specific gene set has a greater mean association with the complex trait than the genes not present in the gene set. Where we identified multiple cell types associated with the same trait, we performed conditional analyses using MAGMA command “--model condition” to evaluate whether it was the same set of genetic variants acting in different cell types or distinct sets of genetic variants in each cell-type suggesting multiple cellular aetiologies. To evaluate the genetic overlap between two traits, each showing an association with the same cell type, we performed GWA conditional analysis with multi-trait-based conditional and joint analysis gca37. We then repeated MAGMA analyses with the GWA summary statistics of one trait adjusted for the second trait and vice versa.

### Cell type-specific PPI network and identification of gene modules

A combined protein-protein interaction network was created based on diverse resources: APID (accessed on September 2023), BIOPLEX (accessed on September 2023), HURI (accessed on September 2023), HitPredict (accessed on September 2023), IntAct (accessed on September 2023), MINT (accessed on September 2023), STRING (accessed on September 2023, restricted to Homo sapiens and physical interactions), CORUM (accessed on September 2023) and Reactome (accessed on September 2023). The combined network consisted of a total of 18,896 genes and 1,228,387 interactions.

A cell-type-specific PPI network was built by extracting PPIs from PPI network between the top 10,000 cell-type-specific genes identified as above. To identify modules of highly interconnected genes in a cell-type-specific PPI network, we employed “cluster_louvain” function in igraph R package^84^. This function implements the multi-level modularity optimisation algorithm, where at each step genes are re-assigned to modules in a greedy way and the process stops when the modularity does not increase in a successive step. Modules with >20 genes are used for further analysis. MAGMA gene set analysis was used to test enrichment in disease-risks across all identified modules.

### Cell proportion test

We tested the change in proportion between cell clusters using a randomization test as implemented in scProportionTest^85^, in which labels from both samples were mixed and 10000 random samples were used to estimate the expected proportion difference. A numeric p value is drawn from the randomization and corrected for multiple testing using FDR. We intersected these results with the p-values derived from a second approach, propeller in the speckle R package^86^, which tests for differences in cell type proportions between multiple experimental conditions using biological replicates in each group. Re-analyses of previously published human and nigral midbrain cellular atlases was performed using the cell type assignments reported by the respective authors. GABAergic neurons were considered separately from DaNs to make such external data more comparable to level 1 annotation in the present study.

### Pseudotime trajectory

The trajectory inference was performed using slingshot^87^ that takes as input UMAP projections and cluster labels defined by Seurat as described above for both gene and transcript level analyses. To define the beginning of the trajectory for pseudotime, we considered the state containing most of the control nuclei. After pseudotime calculation, we compared the pseudotime values between control and PD nuclei and identified the clusters with significant pseudotime difference (adjusted P value < 0.05 by t-test) where appropriate.

Density plots are used to visualise the distributions of clusters (by disease or by subtype) along the trajectory.

### DEGs along pseudotime trajectory

We used switchde^88^, a statistical model and R package, to identify genes and transcripts that exhibit switch-like differential expression along single-cell trajectories. It takes the log normalised TPM matrix of gene expression and the pseudotime values computed for each nuclei by slingshot as described above. Significantly DEGs along a trajectory are identified by q-value < 0.05 and the returned parameter t0 is used as estimated point along the trajectory for a gene turning on or off and used in downstream analyses. We also used tradeSeq^89^ to identify dynamic genes along pseudotime. We employed the within-lineage comparison test (startVsEndTest()) following the vignette (https://www.bioconductor.org/packages/release/bioc/vignettes/tradeSeq/inst/doc/tradeSeq.html).

### Cell communication analysis

Cell communication analysis and visualization were performed using the default setting in CellChat package^90^. All ligand-receptor pairs in cell-cell contact, extracellular matrix (ECM) receptors and secreted signalling were included. Specifically, we focused on the gain or loss of interactions in general cell types, the shift of the outgoing and incoming interactions, and the top signalling pathways altered among cell clusters between PD and control. Significant pairs of ligands-receptors at p-value < 0.05 in cell types of interest are used in downstream analyses.

### Enrichment of MAGMA associated PD risk genes in cell-cell crosstalk

We tested the enrichment of the top 100 MAGMA associated PD risk genes in either ligands or receptors between PD-risk enriched cell types and all other cell types. We then ran 10000 randomisation tests by repeating the enrichment test using less associated PD risk genes (with low ranks in MAGMA gene lists).

### Cell type-specific coexpression and PPI networks

We built cell type-specific coexpression networks by employing BigScale2 R package^91^, a suitable approach for single-cell RNAseq data that computes gene-gene correlations using transformed variables in which expression counts are replaced by *Z*-scores. These *Z*-scores are derived from an unsupervised analysis based on iterative differential expression (DE) between small clusters of cells that also account for different sources of variability in single-cell data. Gene pairs with absolute correlations above 0.8 are retained and further filtered by the presence of a PPI from the PPI network described above.

### List of genes of interest

Dopamine-related genes were extracted from KEGG Pathway. RAS-related genes were extracted from Ingenuity Pathway Analysis.

### TF regulon identification

To identify differentially regulated regulons associated with specific DaN and astrocyte subtypes, we employed SCENIC^92^ with user-recommended settings from the SCENIC vignette (http://htmlpreview.github.io/?https://github.com/aertslab/SCENIC/blob/master/inst/doc/SCENIC_Running.html). Briefly, we took the gene expression matrix for DaNs and astrocytes separately and we ran a correlation analysis using GENIE3 and then ran SCENIC to determine TF modules within these correlations. Using AUCell, we scored nuclei based on regulon activity and plotted these scaled regulon scores on a per-dopaminergic and per-astrocyte subtype basis.

### Meta GWAS PD-T2D vs PD-noT2D

We performed whole genome association studies between Parkinson patients with a history of type 2 diabetes vs no history of type 2 diabetes in three different population cohorts. We considered the Oxford Parkinson Discovery Cohort, including 965 PD with 69 with a history of diabetes; the Tracking cohort, including 1436 PD with 97 history of diabetes; and 5460 participants of the UK Biobank (UK BB) who received a diagnosis of PD since 2023 including with 866 history of diabetes. For UK BB, we used the field in https://biobank.ndph.ox.ac.uk/ukb/field.cgi?id=22009. We ran GWAS using the genetic data and imputation described in^93^ for Tracking and OPDC. We used GLM by using plink2, adjusting for gender, age, and the first two ancestry principal components (for OPDC/Tracking, see^93^; for UKBB, see Table S13). The QQ plot shows a slight deviation from the diagonal line across the entire range, but a strong deviation at the tail, which could suggest potential inflation of p-values due to true signals or systematic bias.

### Transcript quantification

We first ran Mappa *demux* function for demultiplexing and barcode assignment by splitting the data up into barcode-level FASTQ files. We then used Salmon on each barcode-level FASTQ files and set -- numGibbsSamples 30 to generate Gibbs samples or inferential replicates. These replicates are used by Swish function in the R package Fishpond^94^ to test for differential transcript usage, taking account of the uncertainty in transcript abundances. Specifically, it looks for transcripts where the isoform proportions change across condition. Significant DTUs are identified by q-value < 0.05.

### Directed dopaminergic neuron differentiation

DaNs were generated from the iPSCs through a step-by-step differentiation process previously described^95^, with slight modifications. Briefly, 80-90% confluent hiPSCs were cultured on Geltrex-coated plates and maintained in Neurobasal/DMEM-F12/N2/B27-Vit A media containing 200 mM Glutamax and β-mercaptoethanol (N2B27-RA medium). The cells were subjected to dual SMAD inhibition using 10μM SB431542 and 250nM LDN-193189 to promote neuronal differentiation. 500 ng/mL SHH C25II was added from day 0 until day 6 to facilitate midbrain patterning. Differential WNT activation was induced using 0.7μM CHIR99021 until day 4, which was then increased to 5μM CHIR99021 until day 6. On day 6, the cells were treated with accutase and split 1:1 onto culture plates treated with 1X Geltrex, 1:100 fibronectin, and 1:100 laminin. The cells were then maintained in N2B27-RA medium with 5μM CHIR99021 until day 9, which was then replaced with 3μM CHIR99021 on day 10. On day 11, the media was changed to N2B27-RA supplemented with maturation factors: 10 ng/mL BDNF (Brain Derived Neurotrophic Factor), 10 ng/mL GDNF (Glial Derived Neurotrophic Factor), 200μM Ascorbic acid, 1 ng/mL TGFβ, 20μM dbcAMP (dibutyryl Cyclic Adenosine Monophosphate sodium salt), and 10μM DAPT. On day 12, the cells were treated with accutase and plated onto 50 μg/mL poly-D-lysine/50μg/mL laminin-coated culture dishes at a density of 2.5 × 10^5/cm^2^ in DaN differentiation medium until day 21. On day 21, the media was changed to DaN differentiation medium (N2B27+RA medium supplemented with 10 ng/mL BDNF, 10 ng/mL GDNF, 200μM Ascorbic acid, 1 ng/mL TGFβ, 20μM dbcAMP, and 10μM DAPT) supplemented with 1μg/mL laminin and 1X Geltrex until day 45. On day 43 of differentiation, the cells were treated with either 1μM MPP+ iodide (1-Methyl-4-phenylpyridinium) or 1μM 6-OHDA (6-Hydroxydopamine) diluted in DaN differentiation medium for 48 hours, with an untreated condition for each cell line. The company and catalogue numbers of the reagents used are duly tabulated in table S12.

### Immunofluorescence assay and confocal imaging of iPSC cells

After 48 hours of PD inducer treatment, on day 45 of differentiation, the hiPSC-DaNs were washed three times with PBS and fixed with 4% (w/v) paraformaldehyde for 10 minutes at room temperature (RT). The fixed cells were permeabilised with 0.1% Triton in PBS for 15 minutes at RT on a rocker, after which the cells were blocked using 1X blocking buffer containing Triton-X-100, donkey serum, and Bovine Serum Albumin in PBS for 45 minutes at RT. The cells were then incubated with primary antibodies - anti-TH (1:1000), anti-MAP2 (1:2000), and anti-AGTR1 (1:250) diluted in 1X blocking buffer and left overnight at 4 degrees. Following overnight incubation, the cells were washed three times with DPBS and incubated with Alexa Fluor 488/568/647-conjugated donkey anti-sheep/anti-rabbit/anti-chicken respective secondary antibodies (1:2000) in PBS for 1 hour in the dark at RT. The cells were subsequently washed three times with PBS before being visualised under the Leica SP8 confocal microscope. The catalogue numbers and details of antibodies are listed in Table S12.

The untreated and MPP+ Iodide/6-OHDA treated hiPSC-DaNs stained for TH, MAP2, and AGTR1 were imaged using the Leica SP8 confocal microscope at 20X magnification with 2.5X digital zoom.

Sixteen-bit z-stack images were acquired at 1024 × 1024-pixel depth.

### AGTR1 mRNA spatial distribution and quantification using RNAscope in SNpc of PD patients versus healthy controls

The spatial location of AGTR1 mRNA transcripts was confirmed in SNpc in 4 PD patients with Braak stage 5-6 and 5 healthy controls using RNAscope on formalin-fixed paraffin embedded (FFPE) 6-μm thick sections. These were cut from the midbrain block at the level of 3^rd^ nerve from the contralateral hemisphere to frozen punctures used for the snRNAseq analysis (see above). An Hs-AGTR1 probe was used from ACD, Biotechne (Cat number:602841) which has 20ZZ pairs and targets 252-1287 of NM 009585.3. It detects the following variants ENST00000461609.1, ENST00000402260.2, ENST00000418473.7, ENST00000497524.5, ENST00000349243.8, ENST00000404754.2, ENST00000474935.5 and ENST00000475347.5.) *In situ* hybridisation was performed according to the manufacturer’s protocol with positive (Hs-PPIB, cat number: 313901) and negative control (dapB, cat number: 310043) probes on tissue sections and positive control HeLa cells (Cat number: 310045) and human kidney sections, where we expect AGTR1 mRNA to be preponderant (see Human Cell Atlas) The RNAscope 2.5 HD Detection kit with red chromogen was used (Cat number: 322350). Briefly, sections were deparaffinised at 60°C for 1h followed by immersion in 100% xylene (15 min), and then rehydrated through decreasing concentration of industrial demethylated alcohol (IDA) (100%, 90%, 70%), and air dried for 5 min at room temperature (RT). Hydrogen peroxide (H2O2) blocking was performed for 15 min and sections were washed in RNAscope wash buffer. Heat-mediated antigen retrieval was performed at 100°C for 30 min in RNAscope target retrieval buffer, followed by protease plus treatment for 40 min at 40°C and subsequent washes in the RNAscope wash buffer. Sections were then incubated with the Hs-AGTR1 probe for 2 hours at 40°C for hybridisation, washed in buffer and stored overnight at RT in the saline sodium citrate solution (pH=7). The following day, sections were washed and hybridised with 6 amplification steps using the manufacturer’s amplification solutions (1 to 6) in the following order (Amps 1, 3, 5 for 30 min each followed by washes and Amps 2, 4, 6 for 15 min each followed by washes). Chromogenic development of AGTR1 mRNA using Fast Red was performed for 20 min. Sections were subsequently washed in buffer then tap water, counterstained with Harris’ haemaxotylin for 5 min, washed in tap water, dehydrated in increasing concentration of IDA (70%, 90%, 100%), cleared in 100% xylene before coverslipped in distrene-plasticiser-xylene (DPX), dried overnight and scanned using the Aperio scanscope (Leica) at a pixel resolution of 450 nm/pixel.

200 SNpc neurons were assessed in PD and healthy control sections (100 in each group). We used the ACD scoring system to assess the number of transcripts per cell body as follows: score of 0 for no puncta/cell, score of 1 for 1-3 puncta/cell, score of 2 for 4-10 puncta/cell, score of 3 for more than 10 puncta/cell, score of 4 for more than 30 puncta/cell.

### AGTR1 protein quantification using immunohistochemisty in the SNpc of PD patients, ILBD cases and healthy controls

AGTR1 was labelled using immunohistochemistry with a rabbit polyclonal anti-AGTR1 antibody from Proteintech (25343-1-AP, Proteintech, UK) at a 1:500 dilution in 5% fetal bovine serum (FBS) in TBS-T (0.05%, pH=7.6). Briefly, 6-μm thick FFPE sections from SNpc (at the level of 3^rd^ nerve, as above) from 5 PD (Braak stages 5-6), 5 iLBD (Braak stages 1-4), and 6 healthy controls were deparaffinised at 60°C (30 min) and 100% xylene (15 min), rehydrated in graded IDAs (100%, 90%, 70%) and washed in tap water. Similar to RNAscope, we used human kidney sections as positive controls for AGTR1. Heat-mediated antigen retrieval in Tris-EDTA (pH=9) was performed for 30 min and sections were subsequently washed in TBS-T (0.05%), and quenched with 10% H_2_O_2_ for 30 min to block endogenous peroxidase activity. Sections were washed, blocked with 10% FBS in TBS-T for 30 min and incubated overnight with primary antibody at 4°C. On the following day, sections were washed in TBS-T and incubated with secondary antibody-horseradish peroxidase complex in the REAL EnVision detection system (K500711-2, Agilent) for 1 hr at RT. Sections were subsequently washed, and visualised in 3’3-diaminobenzidine (DAB) chromogen. Finally, tissue sections were washed in distilled water, briefly counterstained with Harris’ haematoxylin (~3 min), dehydrated in increasing concentration of IDAs (70%, 90%, 100%, 100%), cleared in xylene (3 × 5 minutes), coverslipped in DPX and dried overnight before brightfield slide scanning as described previously.

AGTR1 coverage was quantified using an artificial neural network (ANN) pixel classifier in QuPath which was trained to detect and differentiate between positive AGTR1 puncta, all nuclei and background. The live prediction feature was utilised throughout the optimisation process to assess the precision of the algorithm with a resolution of 0.5 μm per pixel, no feature normalisation or reduction, and a PCA which retained a variance of zero. An average of 100 annotations per signal type were performed (background, haematoxylin and AGTR1 positive puncta) which achived F1 score of 0.9, suggesting that the training algorithm against the annotations was robust and accurate. After the algorithm was optimised, the SNpc was delineated in each case retaining a similar surface area across all cases and QuPath generated AGTR1 percentage of positive pixels per mm^2^.

### Perineuronal oligodendrocyte labelling and quantification in in the SNpc of PD patients, iLBD cases and healthy controls

Antigen retrieval was in citrate buffer at pH=6. Briefly, 6-μm thick paraffin-embedded sections from the same cases as above i.e. age-matched 4 PD (Braak stages 5-6), 4 iLBD (Braak stages 1-4), and 5 controls were deparaffinised at 60°C (30 min) and 100% xylene (15 min), rehydrated in graded alcohols (100%,90%,70%) and washed in tap water. 1% cresyl violet were filtered overnight and warmed to 60°C prior to staining: sections were placed in solution for 1 min, washed with water for 10 min and differentiated in glacial acetic acid (0.1%), then washed in water, dehydrated in graded alcohol series, cleared in 100% xylene for 15 min, mounted, coverslipped and dried overnight before brightfield slide scanning as described previously. Perineuronal oligodendrocytes were identified by the presence of a small round or oval nucleus, with relatively dense nuclear staining (more chromophilic compared to astrocytic nuclei, whereas macrophages tend to be more rod-shaped) and a narrow unstained rim of cytoplasm^96^. The morphological identification was confirmed by olig2 antibody (Catalogue number: HPA003254) at 1:1000 as previously described. 100 neurons were counted in each case (a total of approximately ~2000 neurons) across all cases: the distance of 5 μm from a neuronal edge was chosen as previously described^96^ and this was manually determined in every neuron using imagescope. The number of oligos/neuron was sampled across SNpc with counting frames placed at random in the region of interest (ROI). The number of perineuronal oligodendrocytes was expressed as the number of cells per neuron. We next calculated the degree of neuronal loss by counting the number of neuromelanised neurons per unit area in the SNpc, which allowed us to determine the neuronal density. We then corrected the mean number of oligodendrocytes per neuron against neuronal densities to obtain a corrected density of perineuronal oligodendrocytes per mm^2^, which reflected the degree of loss.

### Quantification and statistical analysis

Confocal images were analysed using ImageJ software. For AGTR1 expression level analysis, ROI of each cell was first obtained using TH-expressing DaNs. The images acquired at 568nm were separated and analysed for AGTR1 expression in each cell using the saved ROIs, with consistent threshold settings across treatments for each hiPSC-DaN line. Mean intensity from untreated hiPSC-DaNs were normalised to one to facilitate comparisons between conditions. The relative intensity was then calculated and quantified as histograms. Statistical analysis was performed using GraphPad Prism. For statistical significance between the treatments, two-way ANOVA was performed. For comparison of AGTR1 transcripts/cell, protein levels and the density of perineuronal oligodendrocytes between groups, Kruskal-Wallis was used to compare between means with no assumptions about data distributions followed by post-hoc testing corrected for multiple comparisons (Dunn’s comparison). Data are presented as mean ± SEM. Statistical significance was defined as ****p⍰<⍰0.0001.

## Data availability

Raw and processed data to support the findings of this study have been deposited in GEO under accession no.

## Code availability

All code used to generate the main figures of this paper can be found at https://github.com/violavol/Deep_snRNAseq_humanSN_Atlas

## Acknowledgements

The authors would like to sincerely thank all the brain donors for their contribution to this research. We acknowledge the Oxford Brain Bank, supported by the Medical Research Council (MRC), Brains for Dementia Research (BDR) (Alzheimer Society and Alzheimer Research UK), and the National Institute for Health Research (NIHR) Oxford Biomedical Research Centre (BRC), as well as Imperial Brain Bank supported by Parkinson’s UK for supplying tissue. We also thank the computational facilities of the Advance Research Computing at Cardiff (ARCCA) Division, Cardiff University.

## Funding

CW and VV are supported by the UK Dementia Research Institute (MC_PC_17112) which receives its funding from the UK DRI Ltd., funded by the UK Medical Research Council, Alzheimer’s Society and Alzheimer’s Research UK. PS is supported by Parkinson’s UK (project G-2303). LP is supported by the GSK-Institute of Molecular and Computational Medicine, Michael J Fox Foundation, National Institute for Health Research (NIHR) Oxford Biomedical Research Centre (BRC), the National Institute of Health (NIH) and Parkinson Foundation. DAM is supported by the National Institute of Health (NIH). Discovery cohort funding: Parkinson’s UK, and NIHR Oxford BRC. CS, MB, and AK are supported by the UK Dementia Research Institute [award number UK DRI-5209] and a UKRI Future Leaders Fellowship [MR/X032892/1]. CS’s Lectureship position is supported by the Edmond J. Safra Foundation. RWM was supported by the Monument Trust Discovery Award from Parkinson’s UK (J-1403), the BMS/Oxford Alliance and the Michael J Fox Foundation. MTMH received funding/grant support from Parkinson’s UK, Oxford NIHR BRC, University of Oxford, CPT, Lab10X, NIHR, Michael J Fox Foundation, European Platform for Neurodegenerative Disorders (EPND; H2020), GE Healthcare and the PSP Association. SM is funded by a Senior Fellowship from Alzheimer’s Research UK (ARUK-SRF2019A-001).

## Author contributions

Author roles were classified using the Contributor Role Taxonomy (CRediT; https://credit.niso.org/) as follows:

V.V.: formal analysis, software, writing – original draft, writing – review & editing, visualization, data curation, conceptualization;

D.A.M.: method, investigation;

P.S.: method, investigation;

S.G.: method, investigation;

M.R.: investigation;

L.F.C.: investigation;

M.B.: formal analysis, software;

AK.S.: software;

A.Z.: software;

J.M.S.: writing – review & editing;

NN.V.: investigation;

J.M.: investigation;

M.T.M.H: writing – review & editing;

S.M.: writing – review & editing;

R.W.M.: writing – review & editing, project administration, supervision, funding acquisition;

C.S.: formal analysis, writing – review & editing, project administration, supervision, funding acquisition;

L.P.: writing – review & editing, project administration, supervision, funding acquisition;

C.W.: writing – original draft, writing – review & editing, project administration, supervision, conceptualization, funding acquisition;

## Disclosures

Michele T.M. Hu currently receives payment for Advisory Board attendance/consultancy from Helicon, NeuHealth Digital and Manus Neurodynamica. She is an advisory founder and shareholder of NeuHealth Digital Ltd (company number: 14492037), a digital biomarker platform to remotely manage condition progression for Parkinson’s.

## Notes

### Author Declarations

Post-mortem brain tissues from patients with PD, iLBD, and healthy controls were obtained from the Oxford Brain Bank (OBB, University of Oxford, UK) and the Parkinson's UK Brain Bank (Imperial College London, UK), in accordance with approved protocols by the South Central - Oxford C Research Ethics Committee (ref 23/SC/0241) and London Multicentre Research Ethics Committee, respectively. All participants provided informed consent for the brain donation. Both brain banks comply with the requirements of the Human Tissue Act 2004 and the Codes of Practice set by the Human Tissue Authority (HTA licence numbers 12217 for OBB and 12275 for the Imperial).

### Summary of Updates

We have updated Figure 6 and its relative section to include new results on the association between Type 2 Diabetes (T2D) and Parkinson's disease, breaking down T2D into its underlying traits glycemia (as measured through HBA1C) and insulin resistance (as measured through triglyceride levels) which were associated to both AGTR1+ dopaminergic neurons and perineuronal oligodendrocytes.

