## Supplemental Figures for "A deep cellular atlas of the human ventral substantia nigra in Parkinson’s identifies a genetic and molecular overlap with insulin resistance"

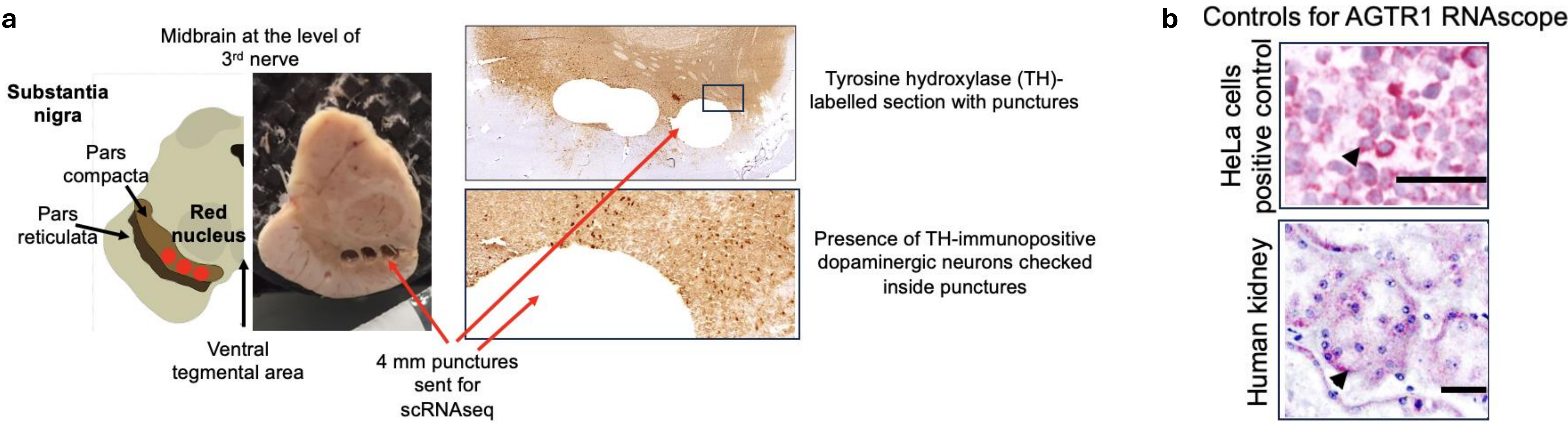

**Figure S1: Sampling method for the ventral substantia nigra pars compacta (SNpc) for sequencing. (a)** 3 × 4 mm punctures were dissected from the ventral tier of the SNpc in frozen midbrain blocks cut at the level of third cranial nerve, obtained from 20 post mortem donors. Two punctures were used for the snRNAseq, and one for a separate ATAC-seq analysis. The midbrain sections were first stained with immunohistochemistry for tyrosine hydroxylase (TH) to anatomically guide the dissection of the punctures (note that in PD cases, the neuromelanin is typically abolished). After the puncture, an additional section was cut and stained with TH to confirm the presence of TH-immunopositive dopaminergic neurons within the puncture sites. **(b)** Positive controls for AGTR1 RNAscope in HeLa cells (fixed under controlled conditions, provided by ACD/Biotech) and control human kidney sections, showing pericellular and peritubular AGTR1 distributions respectively; scalebar = 50 μm.

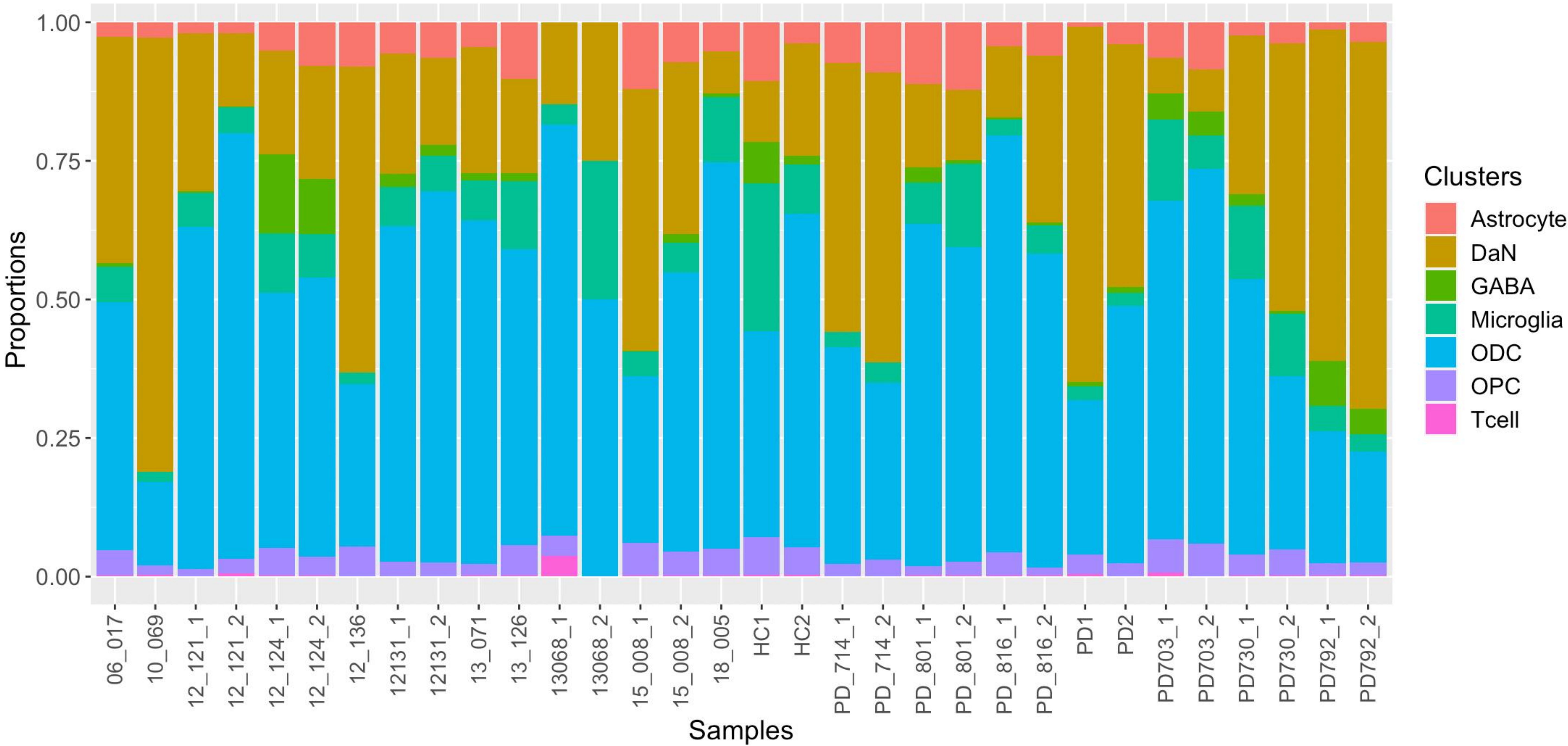

**Figure S2: Cell type proportions across all 32 samples.**

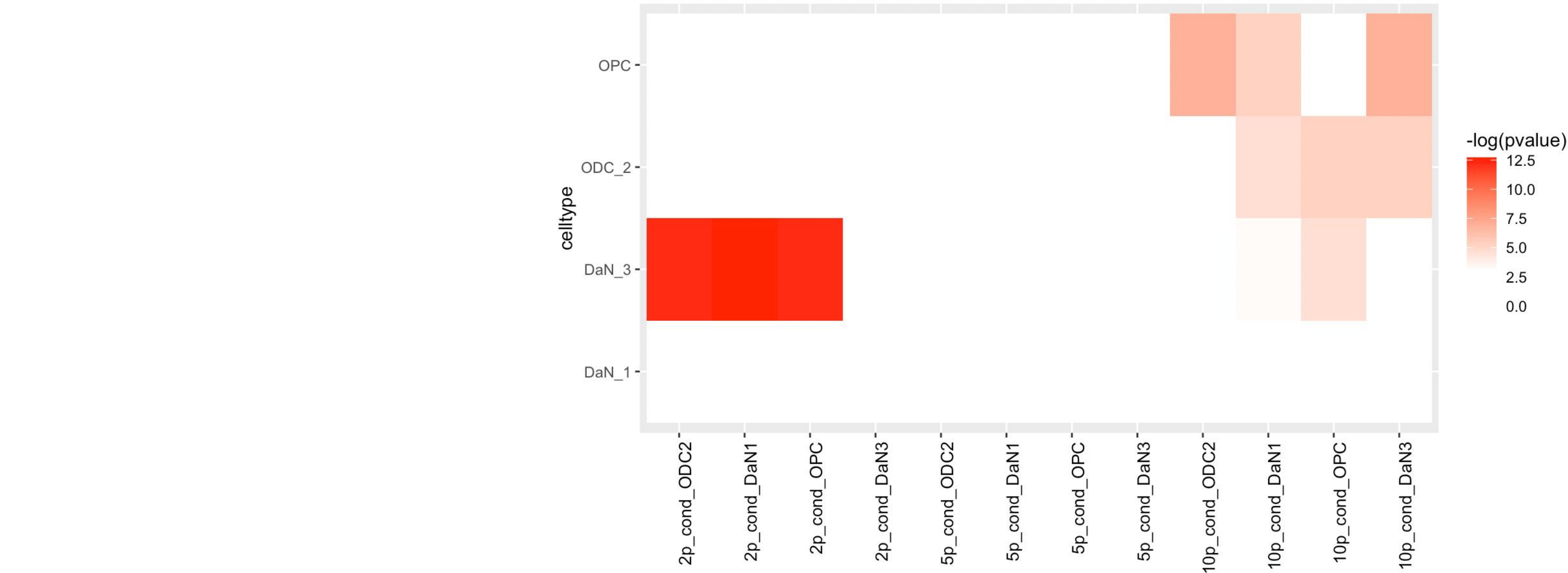

**Figure S3: Conditional analysis of MAGMA PD risk associations.** Heatmap showing $-\log(p\text{-value})$ of conditional associations of PD genetic risk with top 2%, 5% and 10% cell type-specific genes for DaN\_3, DaN\_1, ODC\_2 and OPCs.

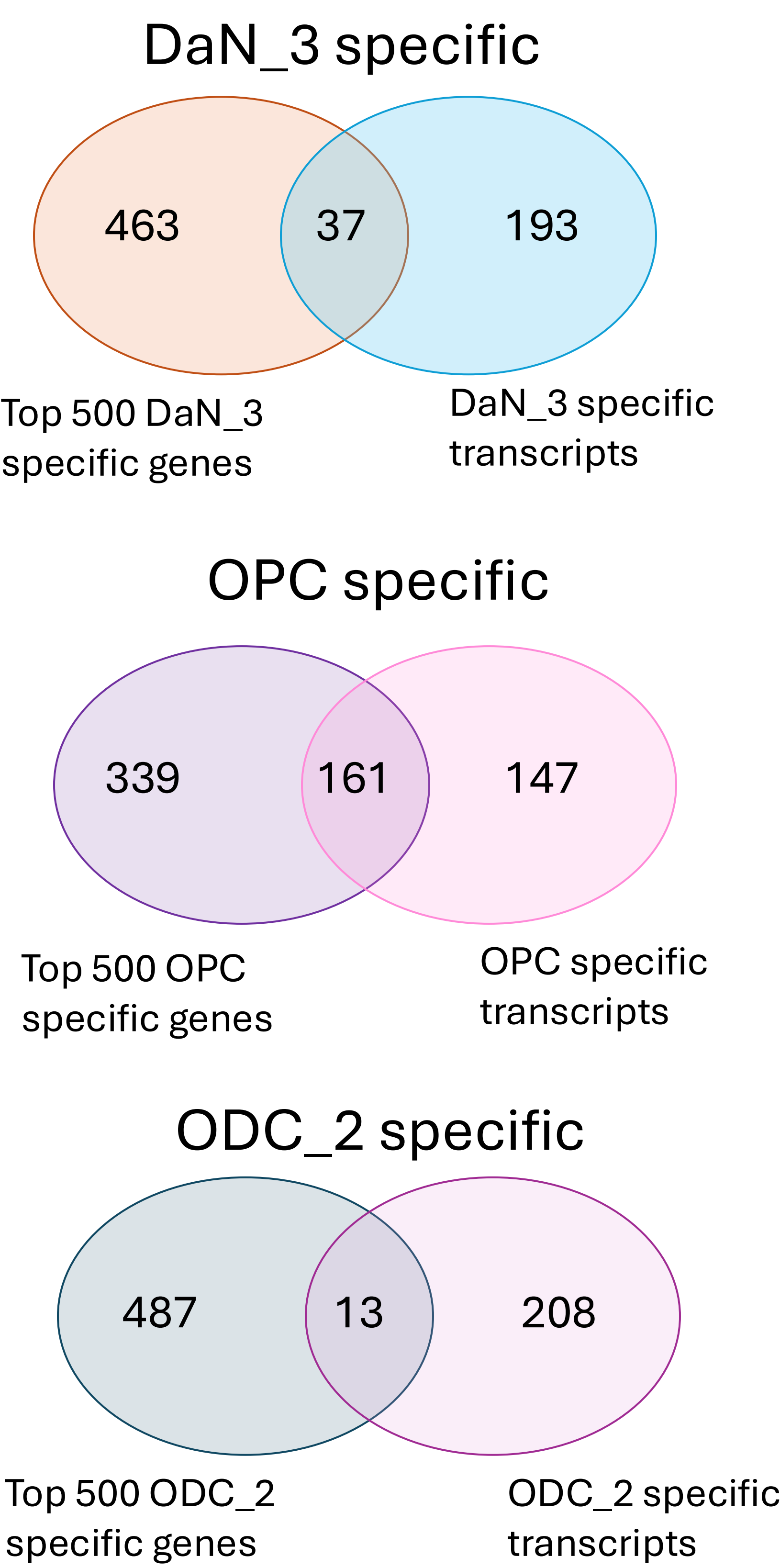

**Figure S4: Overlap between the top 500 PD risk MAGMA-associated DaN\_3, ODC\_2 and OPC-specific and PD risk MAGMA-associated DaN\_3, ODC\_2 and OPC-specific transcripts.**

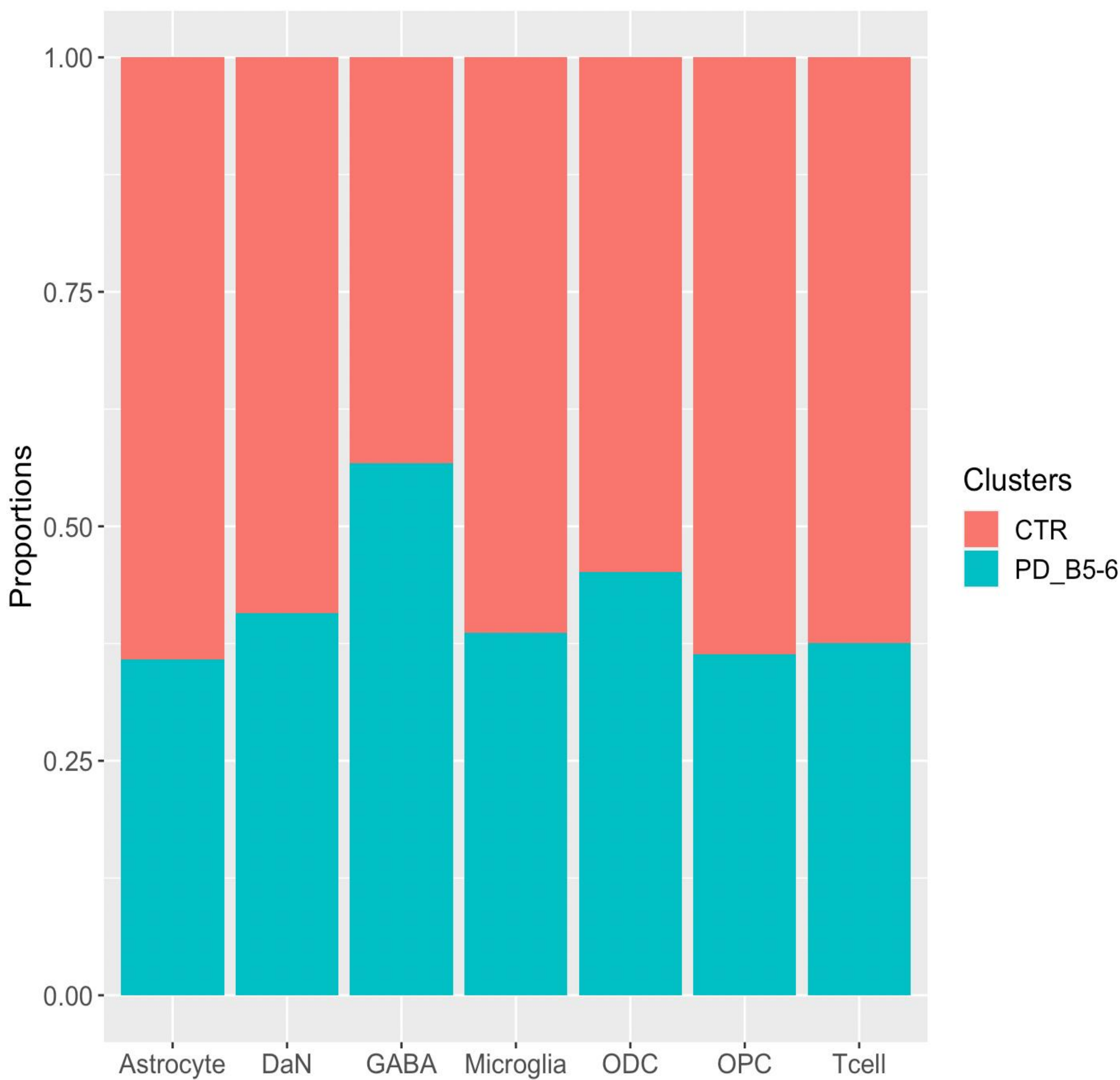

**Figure S5: Cell type proportions in healthy controls and PD Braak stage 5-6 samples.** No significant changes have been found at cell type level 1.

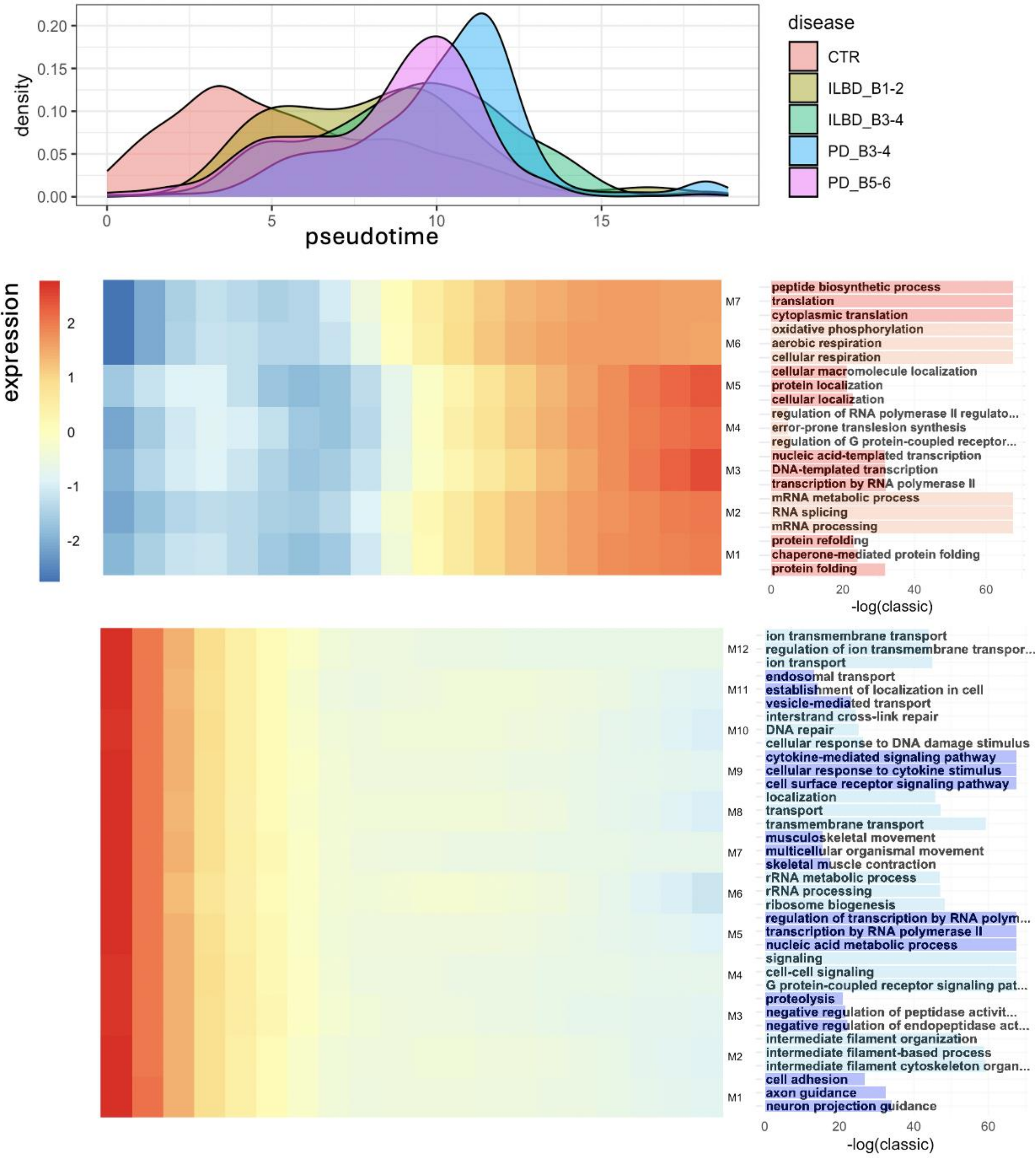

Figure S6: DaN-specific disease progression axis.

Density plot showing pseudotime trajectory for DaN\_0 and DaN\_1 colored by disease condition (top). Heatmap showing expression changes of PPI-based gene modules along the trajectory (bottom). Up and down regulated genes were identified by tradeseq and used to generate the gene modules. Barplot showing $-\log(\text{p-value})$ for the top enriched GO terms by gene module (right).

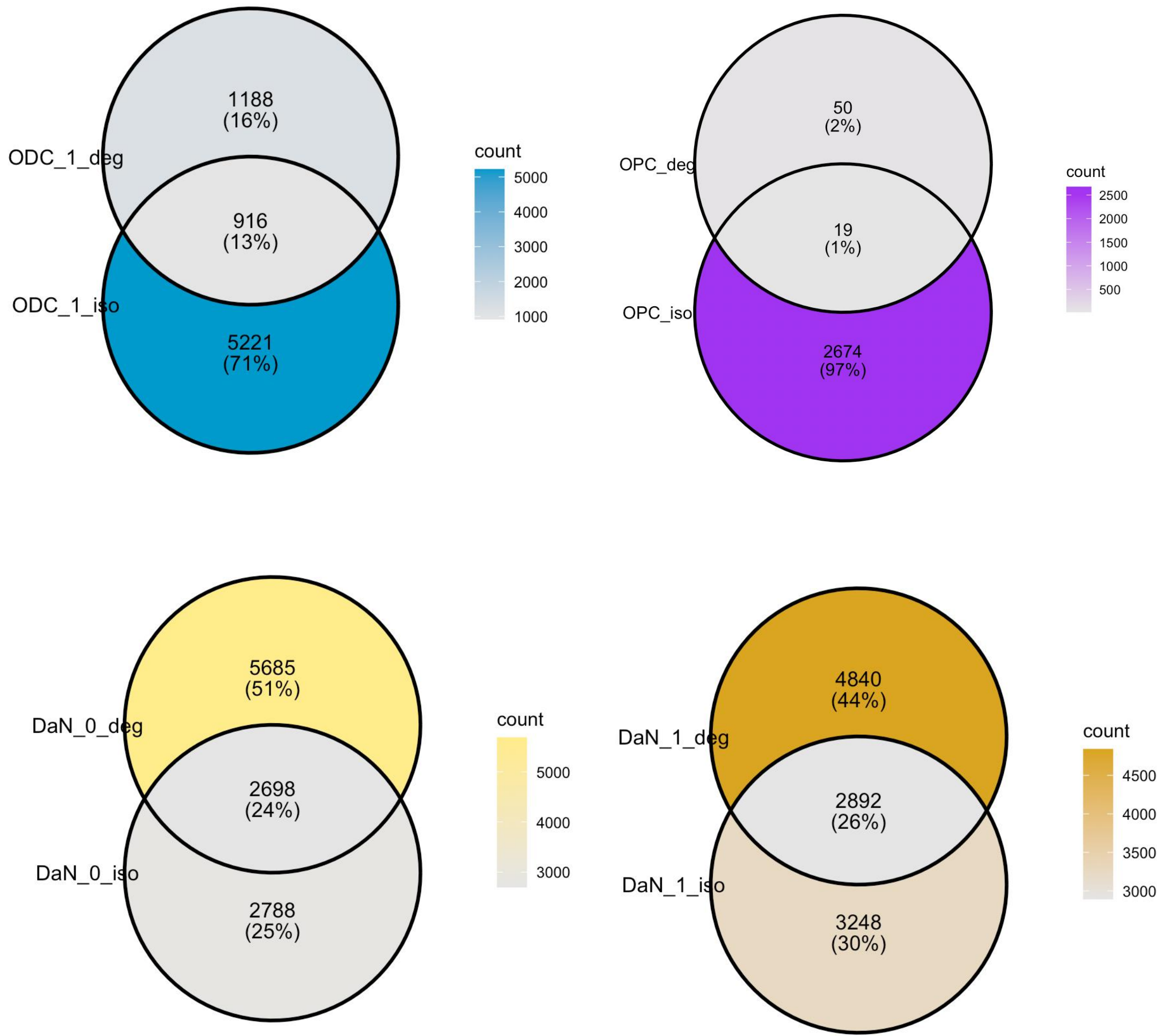

Figure S7: Overlap between DEGs and DTUs between PD\_B5-6 and controls by cell type.

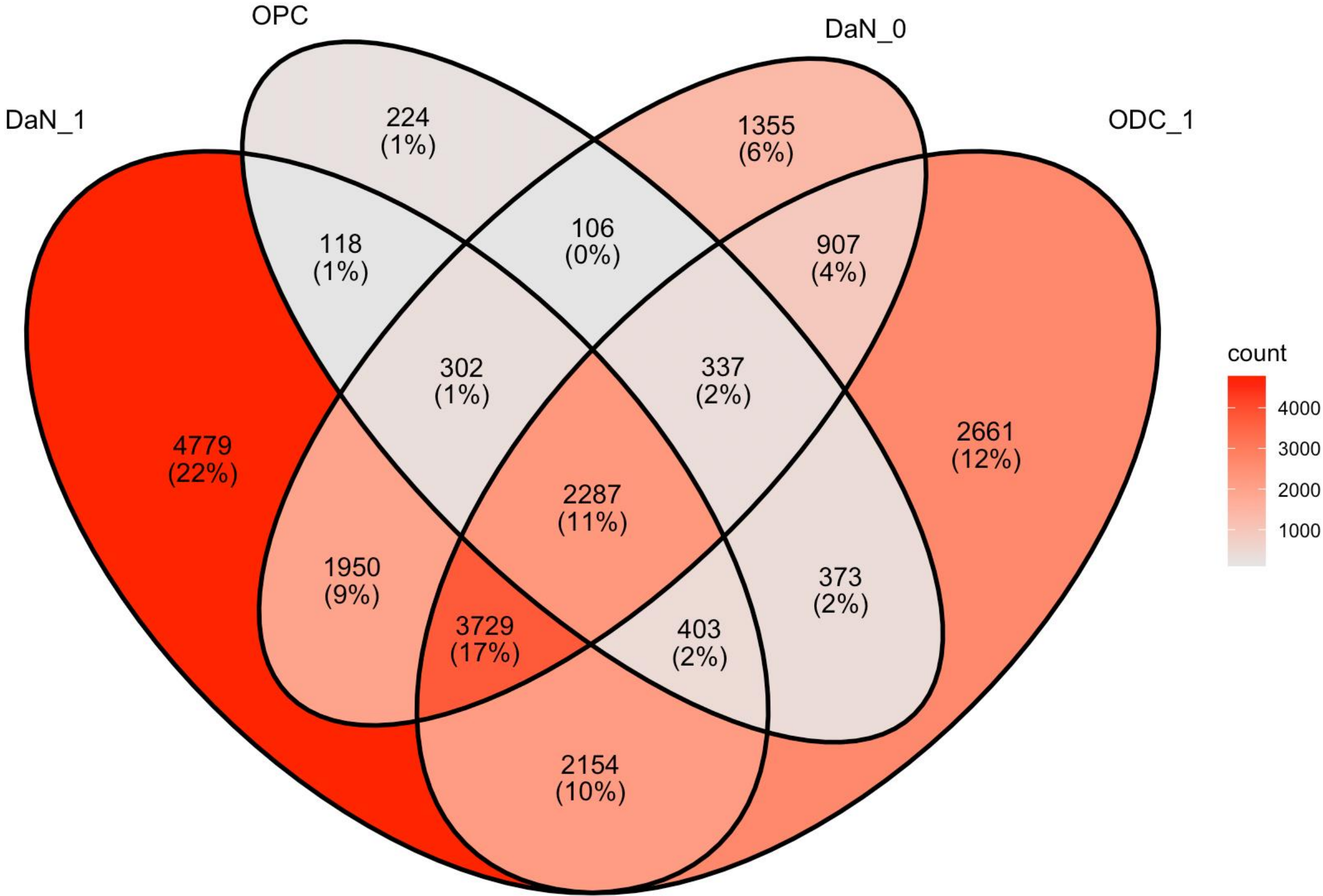

Figure S8: Overlap of DTUs between PD\_B5-6 and controls across cell types.

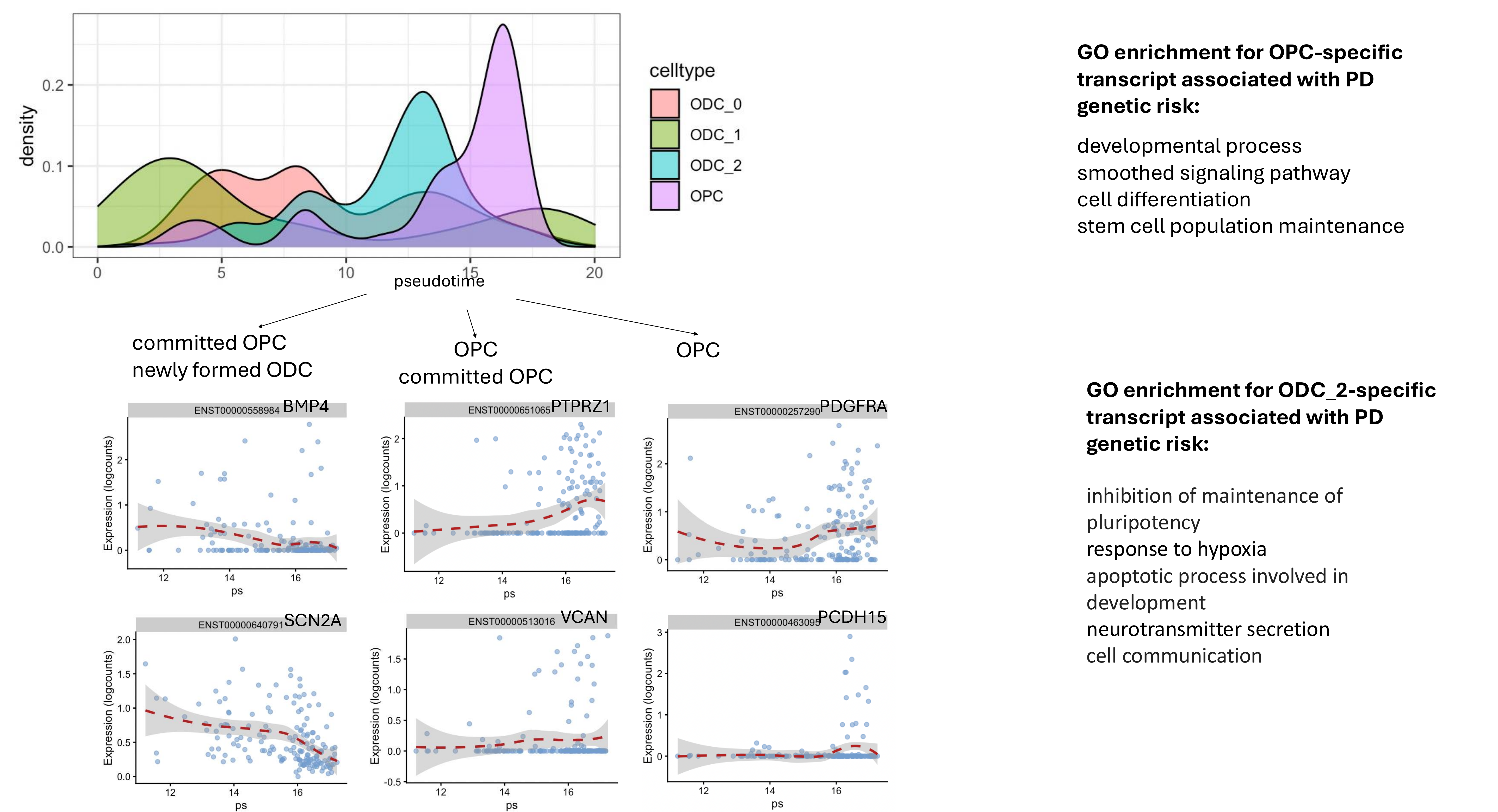

**Figure S9: OPCs to ODCs differentiation at transcript level.** Density plots showing pseudotime trajectory for OPCs differentiation into ODCs computed using isoform expression in controls. Expression of OPC differentiation stage specific genes to confirm correlation of pseudotime trajectory with OPC differentiation process. GO enrichment for OPC and ODC\_2 specific MAGMA associated transcripts with PD genetic risk.

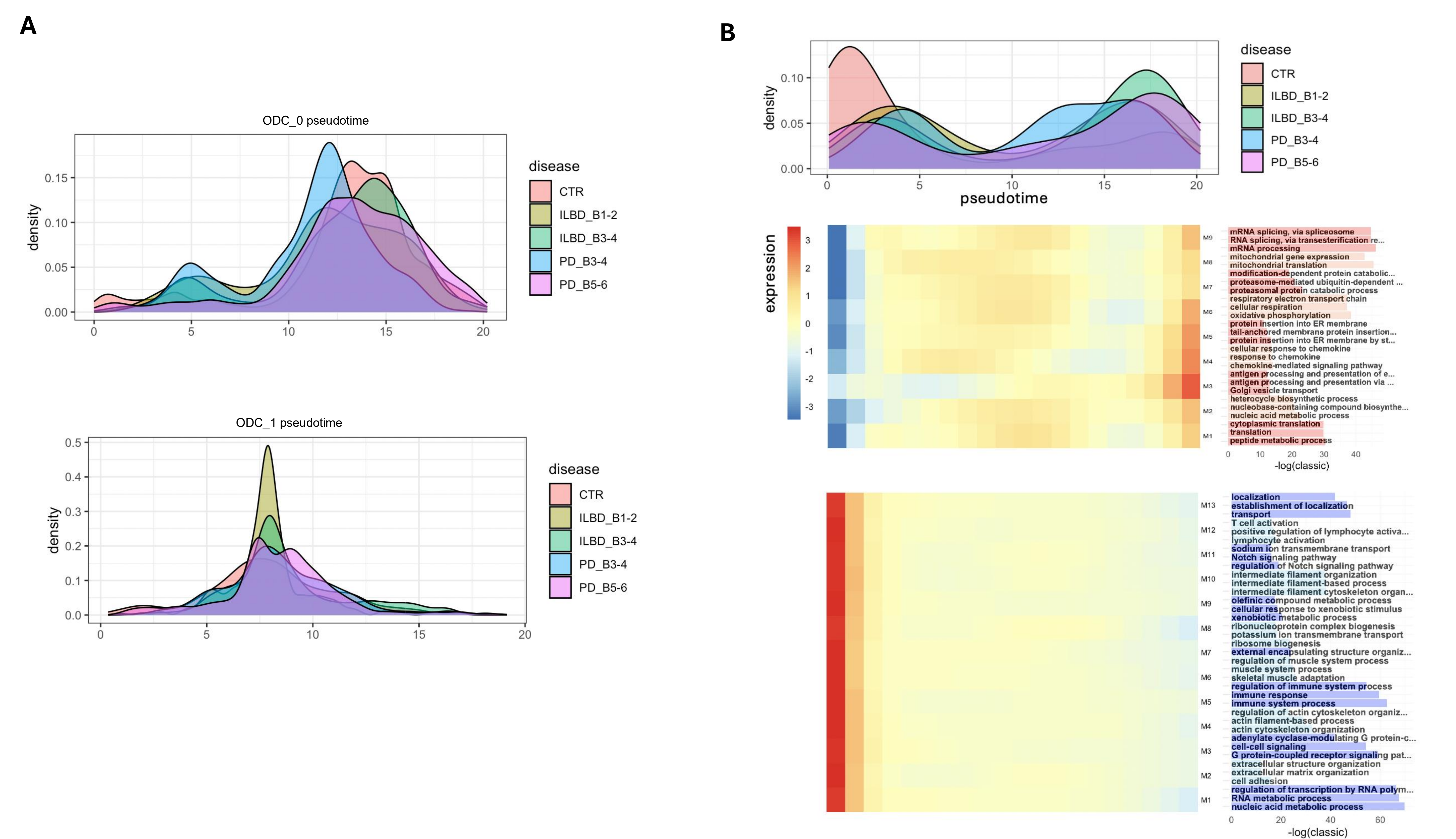

**Figure S10: psODC-2-specific disease progression axis.** (A) Density plots showing pseudotime trajectory for ODC\_0 and ODC\_1 colored by disease condition. (B) Density plot showing pseudotime trajectory for ODC\_2 colored by disease condition (top). Heatmap showing expression changes of PPI-based gene modules along the trajectory (bottom). Up and down regulated genes were identified by tradeseq and used to generate the gene modules. Barplot showing $-\log(p\text{-value})$ for the top enriched GO terms by gene module (right).

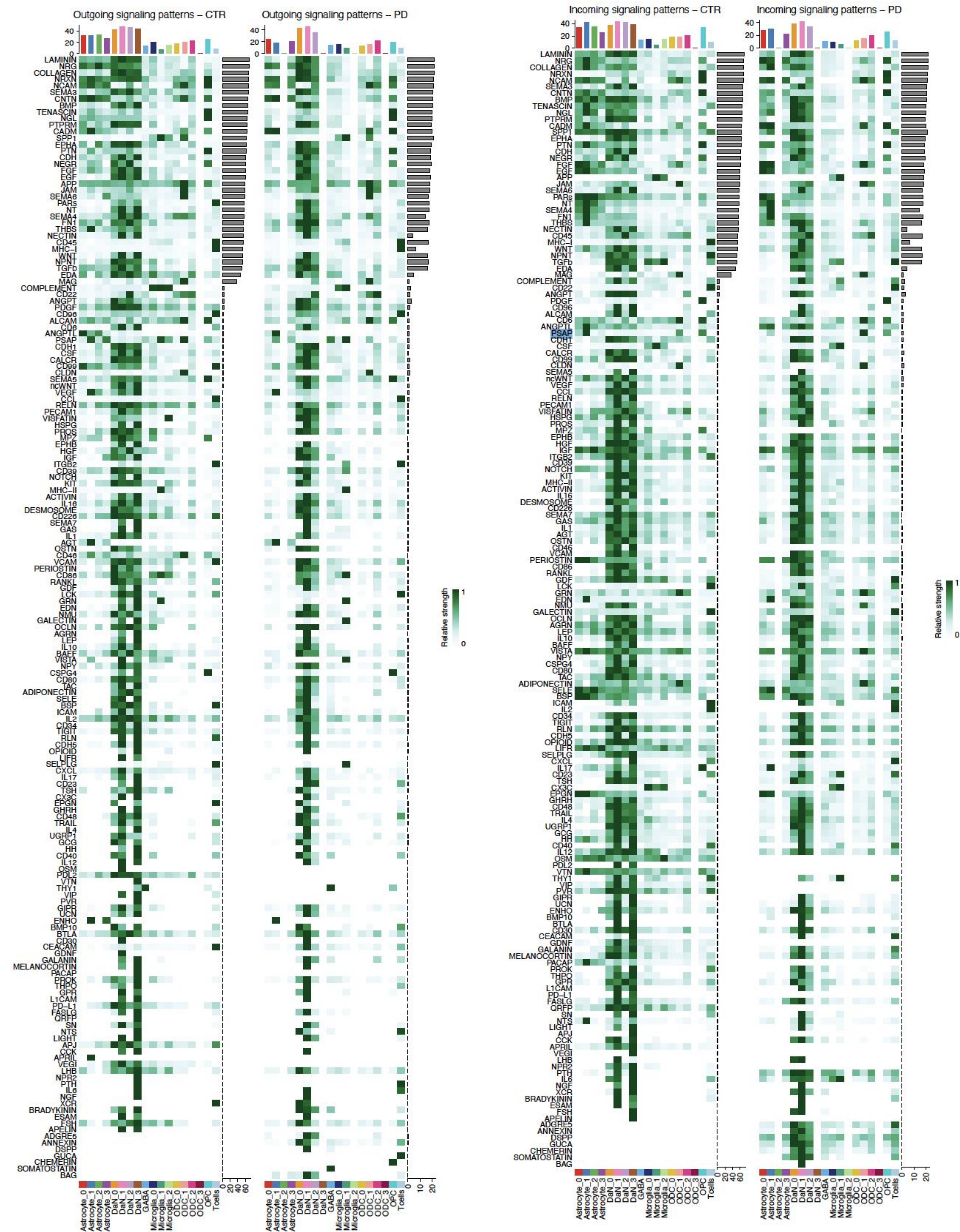

Figure S11: Outgoing and Incoming signaling pathways in controls and PD\_B5-6 from CellChat.

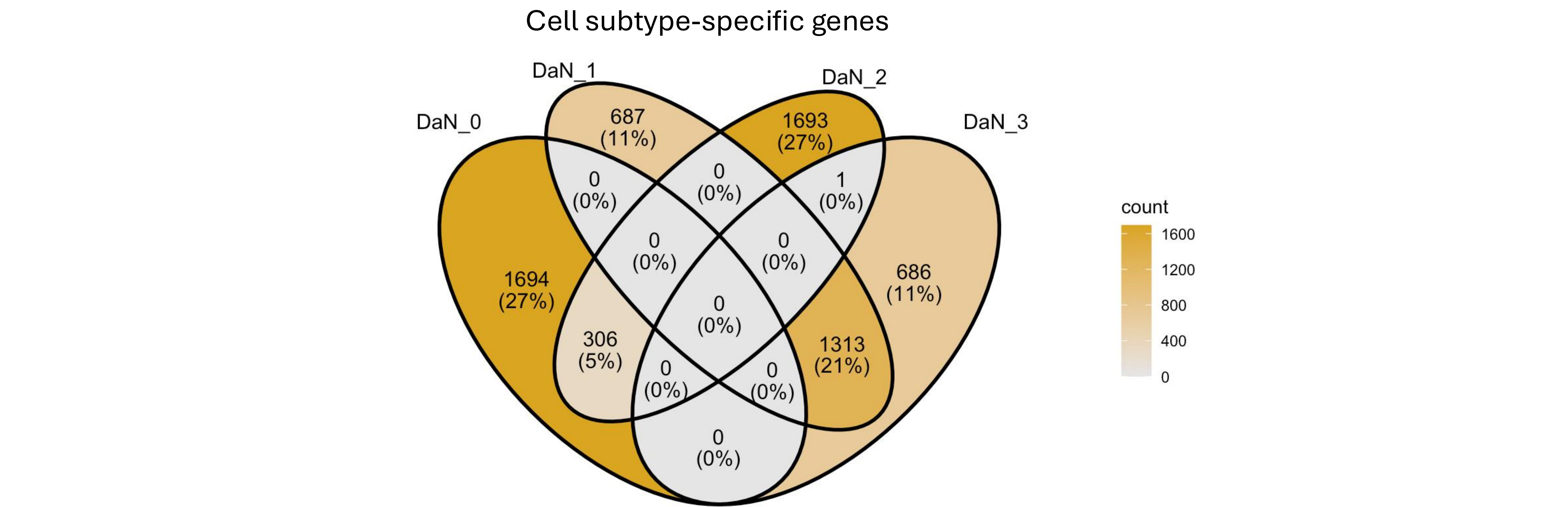

Figure S12: Overlap between top 10% DaN subtype-specific genes.

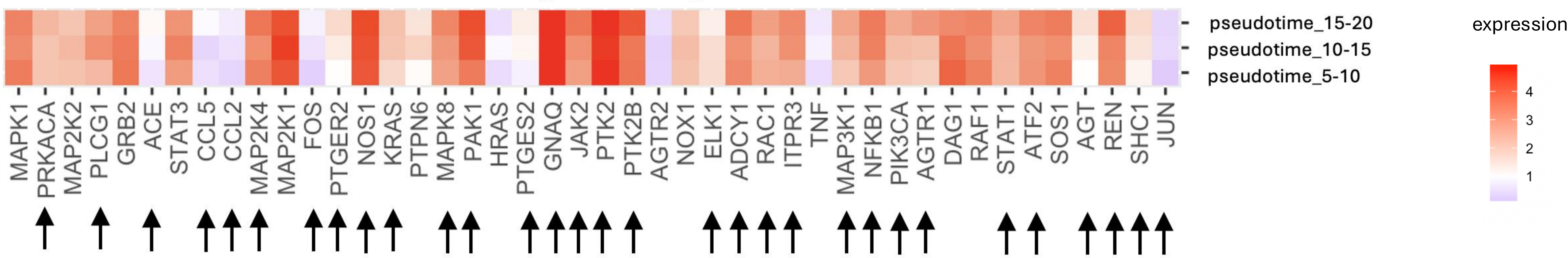

Figure S13: Expression of RAS genes along DaN\_3 transition pseudotime trajectory.

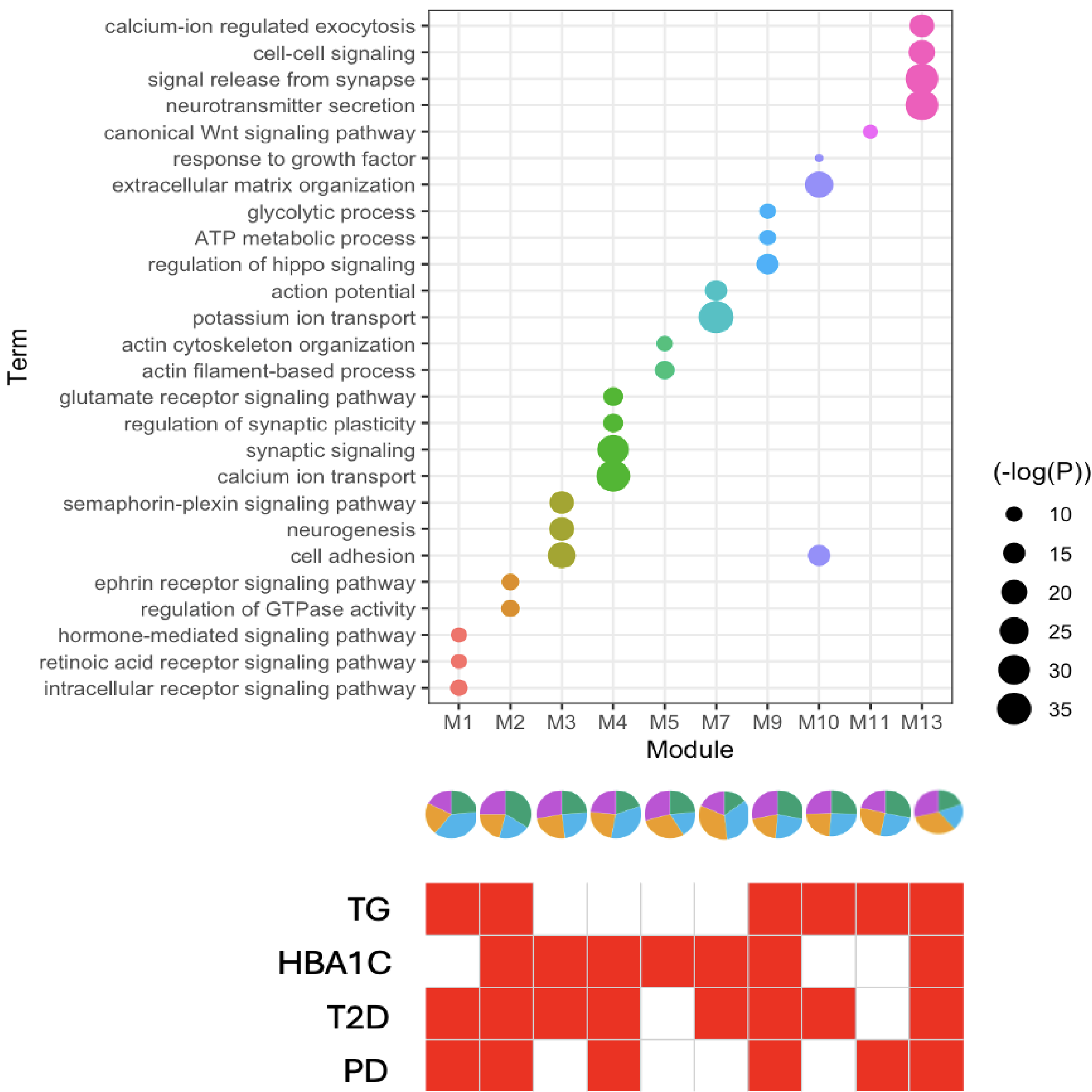

**Figure S14:** Dotplot showing $-\log(\text{p-value})$ for GO enrichment of PD, T2D, HBA1C and TG genetic risk associated genes in ODC\_2-specific PPI modules (top). Pie charts showing the proportions of PD, T2D, HBA1C and TG genetic risk associated genes in any ODC\_2-specific PPI modules (middle). Heatmap showing in red significant PD, T2D, HBA1C and TG genetic risk associations obtained by MAGMA for any ODC\_2-specific PPI modules (bottom).

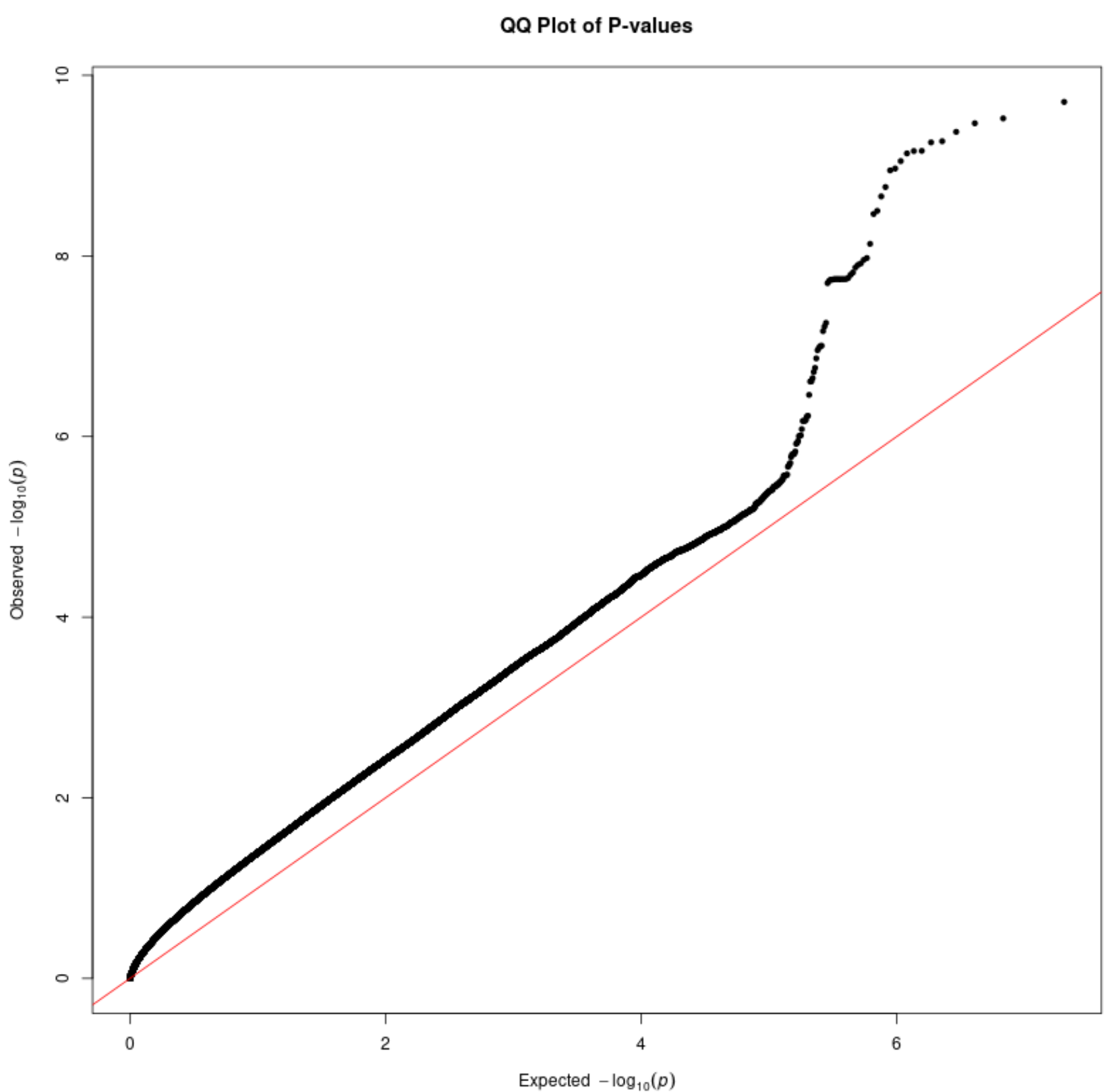

Figure S15: QQ plot of meta GWAS between PD+T2D vs PD-T2D

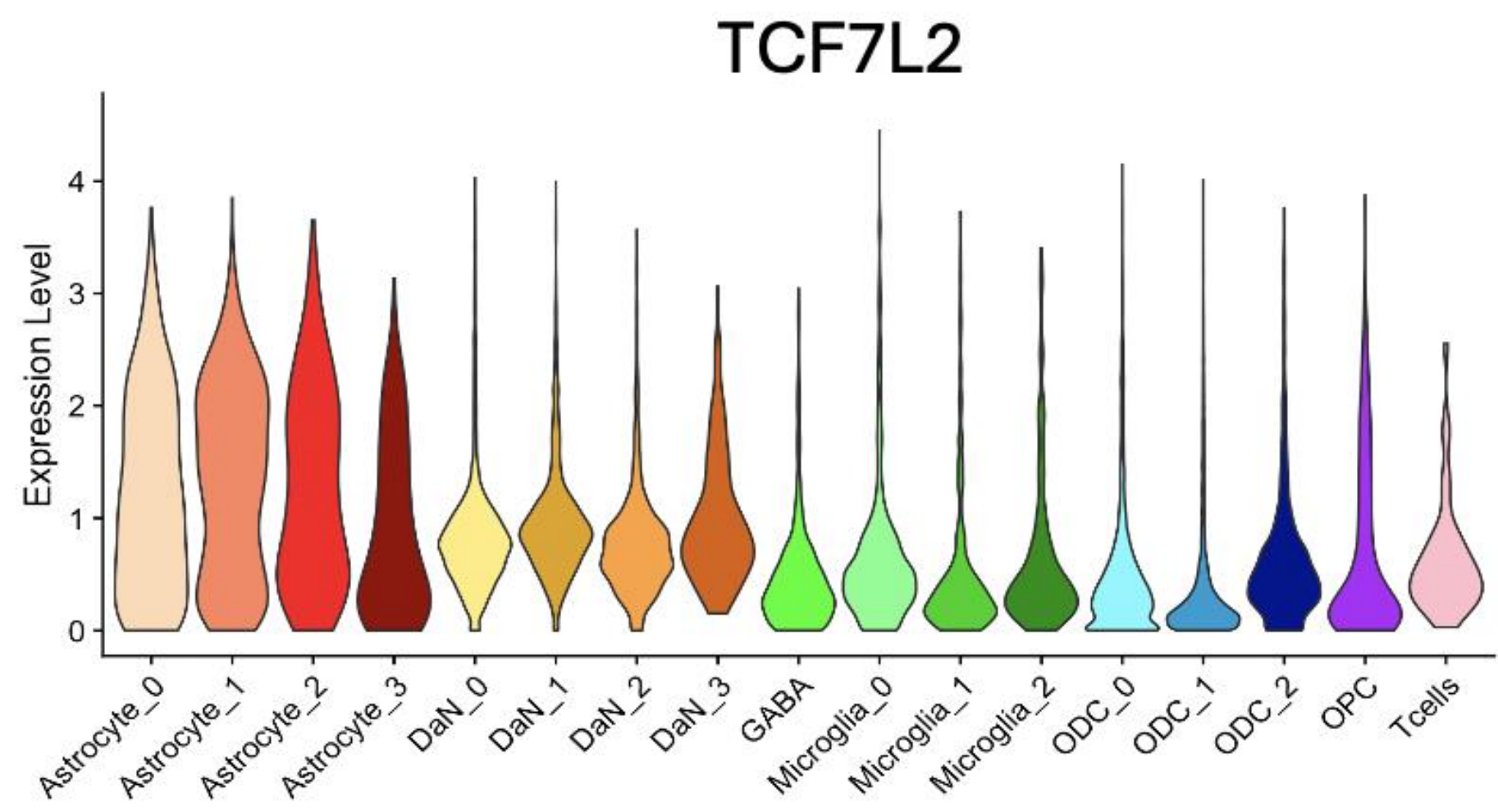

Figure S16: TCF7L2 expression across cell subtypes.

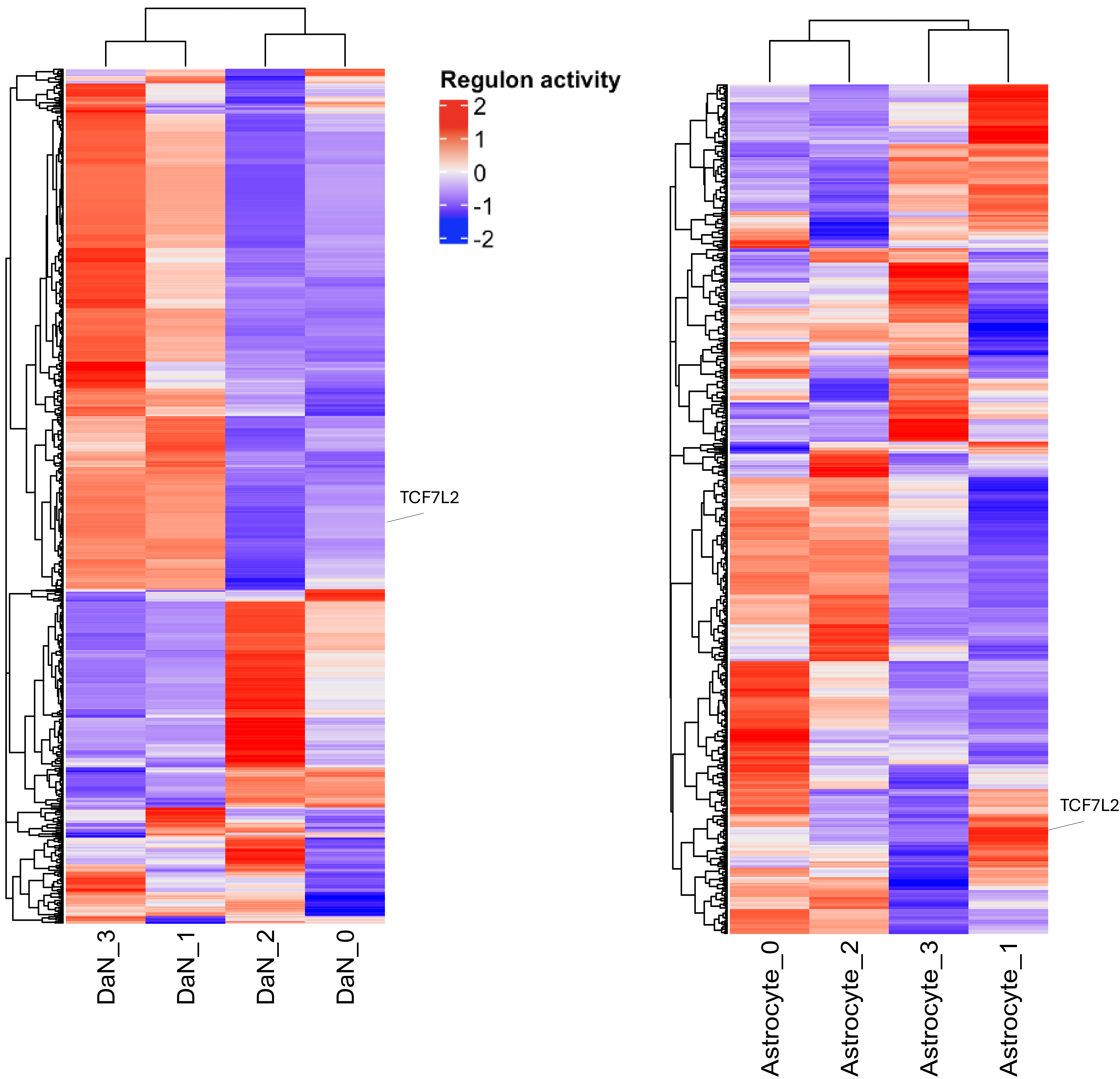

Figure S17: AUC-based regulon activity of transcription factors across DaN and astrocyte subtypes from SCENIC.

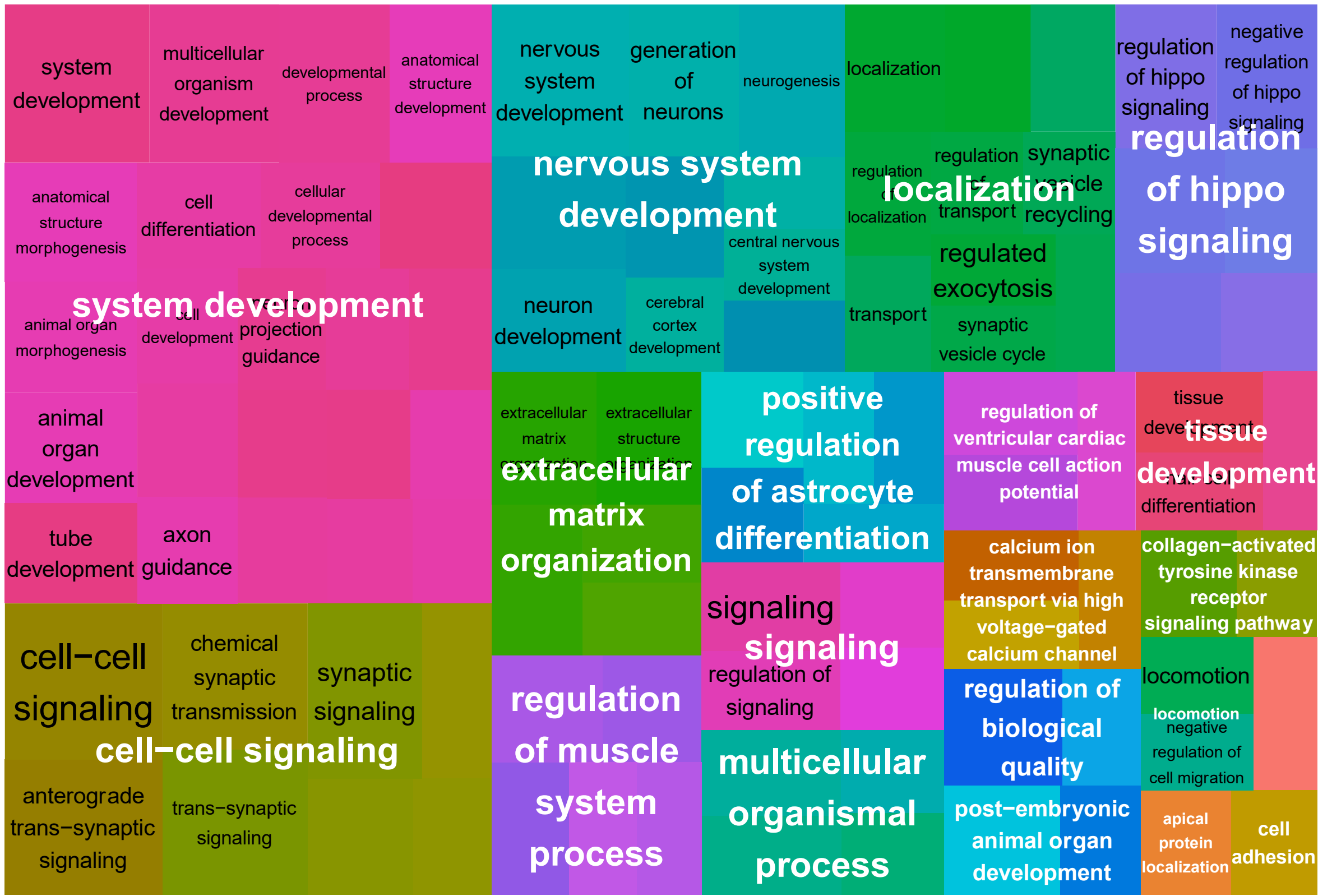

Figure S18: Treemap plots showing enriched GO terms for top 500 MAGMA associated genes with PD+T2D genetic risk

**a**

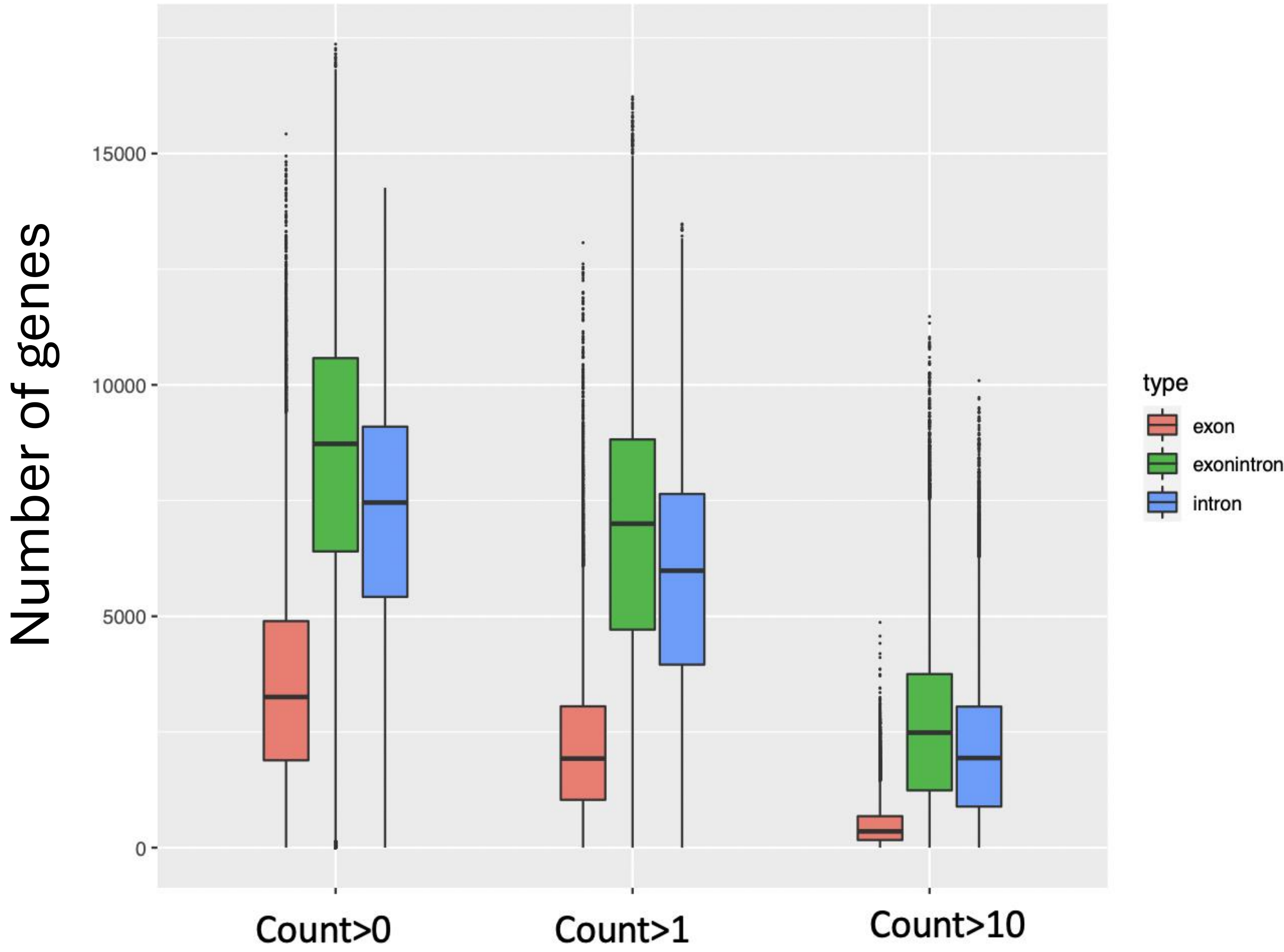

**b**

**Level of expression of cell type specific genes**

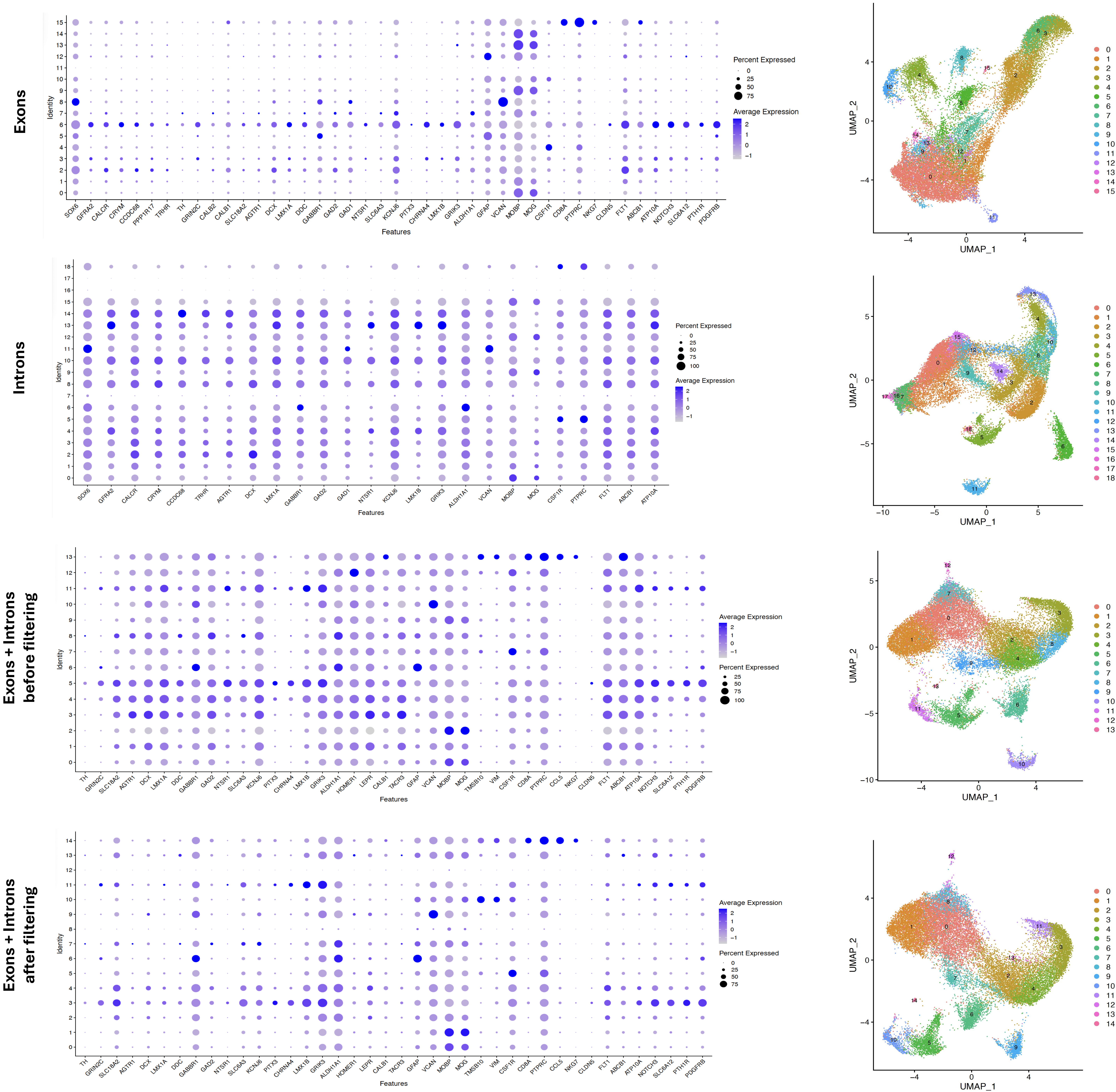
